# Common And Distinct Patterns Of Task-Related Neural Activation Abnormalities In Patients With Remitted And Current Major Depressive Disorder: A Systematic Review And Coordinate-Based Meta-Analysis

**DOI:** 10.1101/2023.03.06.23286814

**Authors:** Zuxing Wang, Peijia Wang, Danmei He, Lin Yang, Hongru Zhu

## Abstract

Whether remitted major depressive disorder (rMDD) and MDD present common or distinct neuropathological mechanisms remains unclear. We performed a meta-analysis of task-related whole-brain functional magnetic resonance imaging (fMRI) using anisotropic effect-size signed differential mapping software to compare brain activation between rMDD/MDD patients and healthy controls (HCs). We included 18 rMDD studies (458 patients and 476 HCs) and 120 MDD studies (3746 patients and 3863 HCs). The results showed that MDD and rMDD patients shared increased neural activation in the right temporal pole and right superior temporal gyrus. Several brain regions, including the right middle temporal gyrus, left inferior parietal, prefrontal cortex, left superior frontal gyrus and striatum, differed significantly between MDD and rMDD. Meta-regression analyses revealed that the percentage of females with MDD was positively associated with brain activity in the right lenticular nucleus/putamen. Our meta-analysis provides useful insights for understanding the potential neuropathology of brain dysfunction in MDD, developing more targeted and efficacious treatment and intervention strategies, and more importantly, providing potential neuroimaging targets for early screening of MDD.

## 1. Introduction

Major depressive disorder (MDD) is a debilitating and common mental disorder that results in considerable social and individual impairment and a substantial burden of the disease overall (Malhi et al., 2020). MDD is a highly recurrent disease with more people undergoing frequent episodes than single episodes (Huang et al., 2019). In recent years, although numerous studies have attempted to pursue more effective approaches for the diagnosis and treatment of MDD, the underlying pathophysiological mechanism remains unclear.

Thanks to better public understanding of MDD and improvements in medical technology, many people suffering from MDD have been able to find complete relief through systematic treatment (Smoski et al., 2013; Wang et al., 2022). Although patients with MDD have shown improvement according to a clinical symptom evaluation, individuals with remitted MDD (rMDD) still demonstrate differences in cognitive function, neuroimaging, and metabolism when compared to healthy controls (HCs) (Moriarty et al., 2022; van Kleef et al., 2022). These comparisons may assist in identifying patients at risk for relapse and improving preventive treatment.

Over the past 30 years, functional magnetic resonance imaging (fMRI) studies have significantly increased our understanding of the neural mechanisms underlying MDD by measuring functional brain activity in MDD patients (Keren et al., 2018; Wang et al., 2015). However, the main problem across experiments of task-fMRI studies in MDD is that the results cannot be replicated to a large extent (Lemke et al., 2022; Li et al., 2022; Trettin et al., 2022). To date, heterogeneity, small sample sizes and differences in analytical methods (e.g., region-of-interest (ROI) or whole-brain analyses) have resulted in inconsistency in data collection and analysis. Previous meta-analyses of MDD and rMDD found many neural activation changes, such as changes in the superior temporal gyrus (STG), prefrontal cortex, insula, striatum, middle temporal gyrus (MTG), superior frontal gyrus and limbic system (Gong et al., 2020; Palmer et al., 2014; Wang et al., 2022). However, it is unclear whether alterations in neural activation can differentiate MDD from rMDD or whether common abnormalities are present in MDD patients in both the current and remitted states. Only a few studies have focused on the comparison of functional neuroimages between rMDD and MDD patients (Ming et al., 2017; Yang et al., 2018). Several meta-analyses have been carried out to analyze the common and distinct patterns of neural activation during various tasks in patients with different mental disorders (Gong et al., 2020; Janiri et al., 2020). However, to the best of our knowledge, no meta-analyses have examined the use of fMRI in assessing common and distinct patterns in brain activity during different tasks between rMDD and MDD patients.

An important question in fMRI studies of MDD patients is whether the abnormally activated brain areas are acquired alterations or inherent to MDD, which is particularly related to the study of the functional neuroanatomical circuits underlying MDD, as growing evidence indicates that the final MDD pathology results from both susceptible and acquired neural alterations (Kambeitz et al., 2017; Thalamuthu et al., 2022). In particular, when exploring between-group alterations in neural activation in MDD patients compared to HCs, it is often unclear whether such functional abnormalities correspond to preexisting vulnerabilities or secondary deficits specific to MDD onset. Acquired deficits may be further separated due to disease-specific neurological abnormalities and brain function changes after MDD remission. The above distinctions may be teased apart by comparing patients with MDD to HCs, rMDD patients to HCs, rMDD patients to MDD patients, and overlap analyses.

This study sought to use a voxel-based meta-analysis to examine brain activation alterations in patients with rMDD and MDD, using data from a large number of task-related studies.

Meta-regressions were performed to explore how age, sex, duration of illness, number of episodes, and severity of depressive symptoms might affect task-related brain activation (Spets and Slotnick, 2021; Yu et al., 2019). Our study not only enables a more precise understanding of the pathophysiological mechanism of MDD but also contributes to the identification of potential biomarkers for prevention and intervention of MDD.

## 2. Methods

### 2.1 Literature Search and Article Inclusion

A comprehensive literature review was performed in the EMBASE, PubMed and Web of Science databases for task-related neuroimaging researches of MDD and rMDD patients published from June 1999 through May 2022. Furthermore, we scoured review articles and references sections of all retrieved studies for additional information. Literature search, study evaluation, and selection were independently performed by two researchers using the following keywords: ‘remitted’, ‘remission’, ‘euthymic’, ‘recovered’, ‘MDD’, ‘unipolar depression’, ‘depressive disorder’, ‘major depressive disorder’, ‘fMRI’, ‘functional magnetic resonance imaging’, and ‘neuroimaging’. The search formulas for above databases are available in the Supplementary Materials. Any discrepancies were resolved by the addition of a third investigator to make a final decision. This study is registered with PROSPERO (registration number CRD42022365170) and we followed the Preferred Reporting Items for Systematic Reviews and Meta-analyses (http://www.prisma-statement.org) guidelines for meta-analyses of observational studies.

Inclusion criteria for studies were as follows: (1) task-related fMRI data of adult rMDD and MDD patients (18–65 years) were compared to those of HCs; (2) results were exhibited using Talairach coordinates or Montreal Neurological Institute (MNI); (3) a whole-brain analysis was conducted to mitigate bias in the regions that were reported; and (4) whenever articles reported findings from the same task paradigm that included samples overlapped, the one with the greatest number of subjects was selected. Studies were not included if (1) task-related fMRI was not used to compare brain activation between rMDD/MDD patients and HCs at the voxel level, (2) they included duplicated datasets, or (3) did not exhibit peak coordinates. A total of 9212 studies on MDD and 1,006 studies on rMDD were identified. After applying the inclusion and exclusion criteria, 18 studies on rMDD and 120 studies on MDD were found to fit the requirements and were included in the meta-analysis (Fig. S1).

### 2.2 Voxel-wise meta-analysis

The anisotropic effect-size signed differential mapping (SDM) software (http://www.sdmproject.com/software/) was used to perform a voxel-wise meta-analysis (Radua et al., 2012) between patients and HCs for rMDD and MDD separately. All the analytical processes in this study were conducted following the SDM tutorial. The SDM used the fitting method of restricted maximum-likelihood estimation of the variance to find an appropriate balance between lack of bias and efficiency (Radua and Mataix-Cols, 2009; Radua et al., 2012). First, peak coordinates of brain activation differences between rMDD/MDD patients and HCs were extracted from each included study. Next, variance and effect-size signed map for each dataset within a gray matter mask according to the peak coordinates and their effect sizes were generated by an anisotropic nonnormalized Gaussian kernel of 20-mm full-width at half-maximum (FWHM) to optimize the specificity and sensitivity of the analysis. Third, a mean map was created by the voxel-wise calculation of the mean of the dataset maps, weighted by the squared root of the sample size of each dataset; with this approach, studies with large sample sizes had a greater weighting of their results. Ultimately, the results were determined to be statistically significant through standard permutation tests and a threshold of p = 0·005 with SDM z score □= □1 and a cluster-level threshold of 10 voxels in the current voxel-wise meta-analysis (Müller et al., 2018).

In 2010, the US National Institute of Mental Health proposed the Research Domain Criteria (RDoC) framework (Insel et al., 2010). The RDoC is the most appropriate framework for classifying the array of criteria used in research and enables a more comprehensive interpretation of results in terms of dysfunction in clearly defined brain activity processes. According to the RDoC, task-related fMRI studies involving comparisons between HCs and MDD/rMDD patients can be divided into five domains (as classified in Table S1 of the Supplementary Materials).

The results from all tasks and cognitive domains used in the primary experiments were pooled together to generate the modeled activation maps. The RDoC framework (Insel et al., 2010) classifies neurocognitive tasks based on their presumed association with known brain circuits in a principled way. Although there is some relationship between tasks and brain areas, it is not a one-to-one correspondence. The association between brain function and structure has been described both as pluripotent (one□to □many) and degenerate (many □to □one). This means that any given task paradigm can engage brain areas outside these predicted by the cognitive components attributed to that specific task, while a single brain region may be excited by different tasks that may underlying a distinct cognitive mechanism (Pessoa, 2014; Price and Friston, 2005).

Next, to assess the effects between rMDD and MDD patients, a linear meta-regression model was conducted to analyze group comparisons. The results of this group comparison were masked by the findings of an initial model that included both the rMDD and MDD groups; thus, the evaluation only included the main group effect. Only differences that survived a voxel-level threshold of p < 0·005, a cluster-level threshold of k ≥ 10 voxels, and an SDM-Z value threshold of 1 were reported. These threshold corresponded to p < 0·05 corrected for multiple comparisons in a meta-analysis (Norman et al., 2016; Radua et al., 2014, 2010; Schulze et al., 2019).

Finally, the overlap of neural activation differences between rMDD and MDD patients via the combination of thresholded meta-analytic result maps was investigated. This multimodal analysis function of the SDM statistical package enabled us to identify brain areas in which both rMDD and MDD patients share characteristics that differ from those of HCs ((rMDD vs. HCs) versus (MDD vs. HCs)), while taking into account error in the estimation of the magnitude of these differences. The default parameters of SDM were used (peak p < 0·00025, cluster size of >10) (Radua et al., 2010).

### 2.3 Heterogeneity and Publication Bias

A random effects model with *I*^2^ statistics was used to conduct a heterogeneity analysis in order to explore unexplained variability among studies. The *I*^2^ statistic was utilized to evaluate the percentage of variation due to heterogeneity other than chance. Extreme heterogeneity corresponds to *I*^2^ values of 75–100%, large heterogeneity corresponds to *I*^2^ values of 50–75%, moderate heterogeneity corresponds to *I*^2^ values of 25–50%, and low heterogeneity corresponds to *I*^2^ values of 0–25%. Using the default SDM kernel size and thresholds (FWHM = 20 mm, p = 0·005, uncorrected for the false discovery rate (FDR), peak height Z = 1, cluster extent = 10 voxels), we obtained heterogeneous brain regions (Amad et al., 2019; Gong et al., 2020; Radua et al., 2014). Egger’s test was used to evaluate underlying publication bias, with a p value less than 0·05 indicating significant (Egger et al., 1997).

### 2.4 Reliability and Meta-regressions

To assess the robustness of the rMDD and MDD results, a jackknife sensitivity analysis was conducted which entailed excluding one study from the analyses (Radua and Mataix-Cols, 2009). Finally, we used meta-regression analyses, which are based on simple linear regression models to estimate the effect of age, female percentage, the number of depressive episodes, the severity of depressive symptoms and duration of illness on the effect sizes found in the rMDD and MDD meta-analyses. Meta-regression analyses were conducted only when brain areas of rMDD or MDD patients showed significant hypoactivation or hyperactivation compared with those of HCs.

## 3. Results

### 3.1. Study Characteristics

A total of 18 studies and 24 experiments were included, with observations from 440 rMDD patients and 456 HCs. Supplementary. The included studies reported neural activation differences between 3079 MDD patients and 3214 HCs using data from 120 studies which included 147 experiments. Detailed demographic characteristics and study data for both MDD and rMDD can be found in Supplementary Table S2 and S3.

### 3.2. Neural Activation Differences in the Main Meta-analysis

#### 3.2.1. rMDD versus HCs

As shown in Fig. 1A and Table 1, compared with HCs, patients with rMDD indicated significantly increased neural activation in the right inferior network extending to the MTG and STG, right striatum, left median cingulate, and right anterior thalamic projections. They also revealed decreased activation in the right MTG, left superior frontal gyrus (SFG), and left cuneus cortex relative to that of HCs. These brain regions show low between-study heterogeneity. Egger’s tests of publication bias were nonsignificant (all p values > 0·05) except in the right inferior network (*p* = 0·007). Jackknife sensitivity analyses revealed that abnormal neural activation in the right inferior network, SFG, and right MTG were found in all 24 analyses. The other activated or deactivated brain areas remained replicable, as they were significant in at least 16/24 of the experiments (Table 1).

**Figure.**
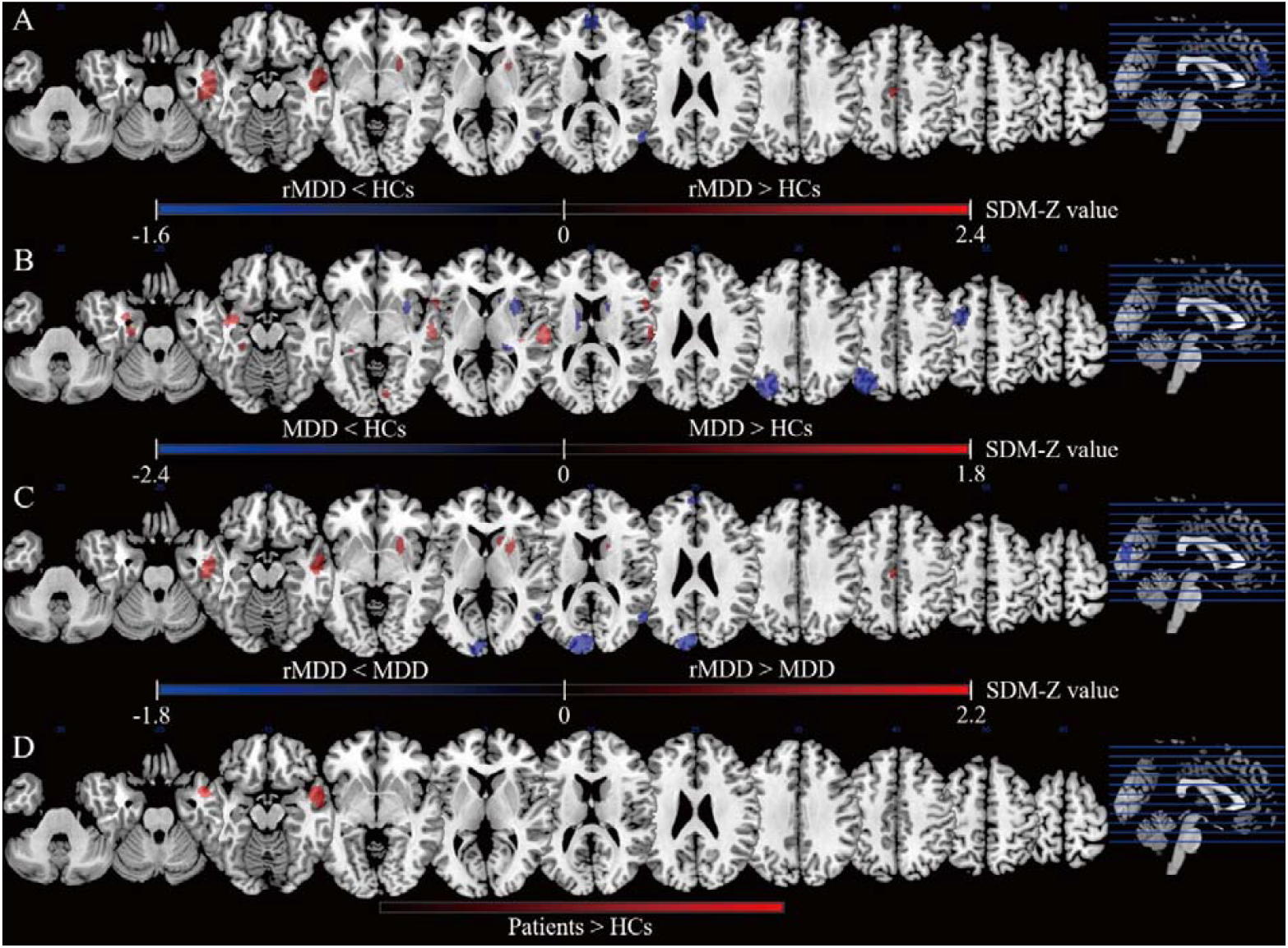
Brain regions showed significant task-related neural activation differences between groups based on the meta-analyses. Meta-analyses results regarding A) task-related neural activation difference between rMDD and HCs, B) task-related neural activation difference MDD and HCs, C) task-related neural activation difference between rMDD and MDD (vs. HCs), as well as D) conjunction of rMDD and MDD (vs. HCs). Areas with decreased task-related neural activation value are displayed in blue, and areas with increased task-related neural activation value are displayed in red. The color bar indicates the maximum and minimum SDM-Z values. Abbreviation: HCs, healthy controls; MDD, major depressive disorder; rMDD, remitted major depressive disorder; SDM seed-based *d* mapping.

**Table 1.** Clusters showing differences between rMDD and HC.

| Local maximum region | Peak MNI coordinate<br>(x, y, z) | SDM-Z | p | Cluster size<br>/ voxels | Breakdown (Number of voxels) | Jackknife<br>sensitivity | Heterogeneity<br>$I^2$ | Publish bias<br>Egger's tests |
| --- | --- | --- | --- | --- | --- | --- | --- | --- |
| rMDD > HC |  |  |  |  |  |  |  |  |
| Right inferior network,<br>inferior longitudinal<br>fasciculus | 48, -14, -22 | 2.436 | 0.000015497 | 862 | Right middle temporal gyrus, BA 21 (257)<br>Right inferior network, inferior longitudinal<br>fasciculus (230)<br>Right inferior temporal gyrus, BA 20 (199)<br>Right temporal pole, middle temporal gyrus,<br>BA 21 (50)<br>Right superior temporal gyrus, BA 21 (36)<br>Right fusiform gyrus, BA 20 (31) | 24/24 | 0.30% | 0.007 |
| Right striatum | 24, 12, 2 | 1.818 | 0.000500619 | 122 | Right striatum (87)<br>Right lenticular nucleus, putamen, BA 48 (24) | 23/24 | 0.10% | 0.228 |
| Left median cingulate /<br>paracingulate gyrus,<br>BA 23 | -4, -16, 44 | 1.562 | 0.002420425 | 53 | Left median cingulate / paracingulate gyrus,<br>BA 23 (52) | 22/24 | 0.0% | 0.688 |
| Right anterior thalamic<br>projections | 18, 14, 10 | 1.527 | 0.002941668 | 26 | Right anterior thalamic projections (25) | 23/24 | 0.0% | 0.833 |
| rMDD < HC |  |  |  |  |  |  |  |  |
| Left superior frontal<br>gyrus | 2, 58, 28 | -1.663 | 0.000206411 | 670 | Left superior frontal gyrus, BA 10 (330)<br>Right superior frontal gyrus, BA 10 (191)<br>Corpus callosum (47)<br>Left superior frontal gyrus, BA 32 (45)<br>Right superior frontal gyrus, BA 32 (31) | 24/24 | 25.0% | 0.126 |
| Right middle temporal<br>gyrus, BA 39 | 52, -68, 14 | -1.338 | 0.001641154 | 155 | Right middle temporal gyrus, BA 37 (119)<br>Right middle temporal gyrus, BA 39 (28) | 24/24 | 11.9% | 0.698 |
| Left cuneus cortex, BA<br>18 | -6, -84, 28 | -1.217 | 0.003602266 | 11 | Left cuneus cortex, BA 18 (11) | 16/24 | 2.1% | 0.897 |

rMDD, remitted major depressive disorder; HC, healthy control; BA, Brodman areas; MNI, montreal neurological institute.

#### 3.2.2. MDD versus HCs

As presented in Fig. 1B and Table 2, the meta-analytic brain map revealed both increased and decreased neural activation in MDD patients compared with HCs. MDD patients showed increased neural activation in the bilateral STG, left insula, left hippocampus, left median network/cingulum (extending to the hippocampus), bilateral inferior frontal gyrus (IFG), right inferior network, and right middle frontal gyrus and decreased neural activation in the left inferior parietal gyrus (extending to the occipital cortex), right lenticular nucleus/putamen, left middle frontal gyrus (including the dorsolateral prefrontal cortex), right thalamus, and bilateral caudate nucleus. These brain areas did not show obvious between-study heterogeneity. Egger’s tests of publication bias were nonsignificant (all p values > 0·05) except for the right inferior network (*p* values = 0·038). The jackknife sensitivity analysis showed that in MDD patients, the most robust brain area patterns were increases in neural activation in the right STG and left insula and a decrease in neural activation in the left inferior parietal gyrus, as they were significant in all 147 analyses. The increased or decreased neural activation in the other brain regions remained replicable, as they were significant in at least 126/147 of the experiments (Table 2).

**Table 2.**
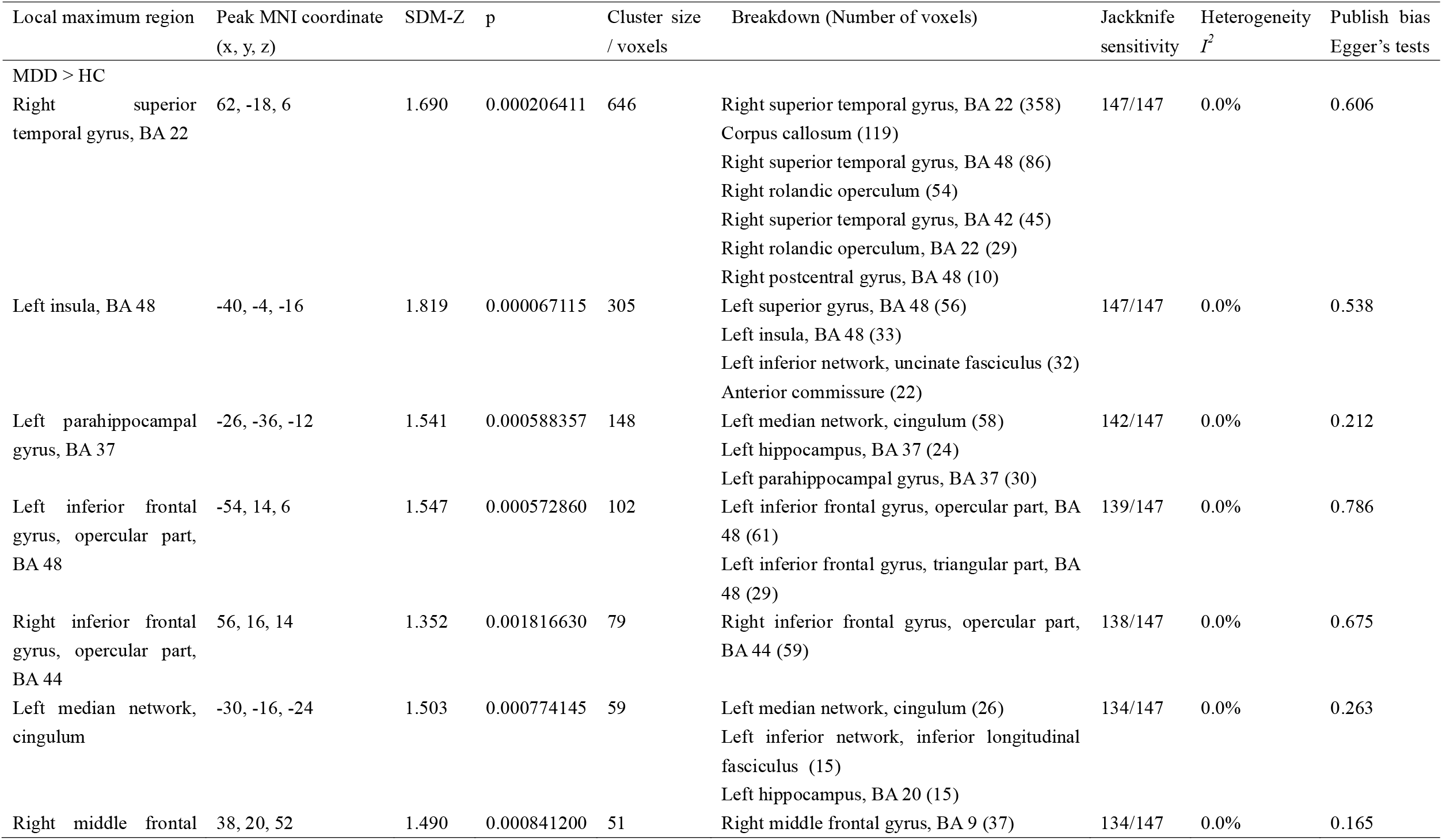
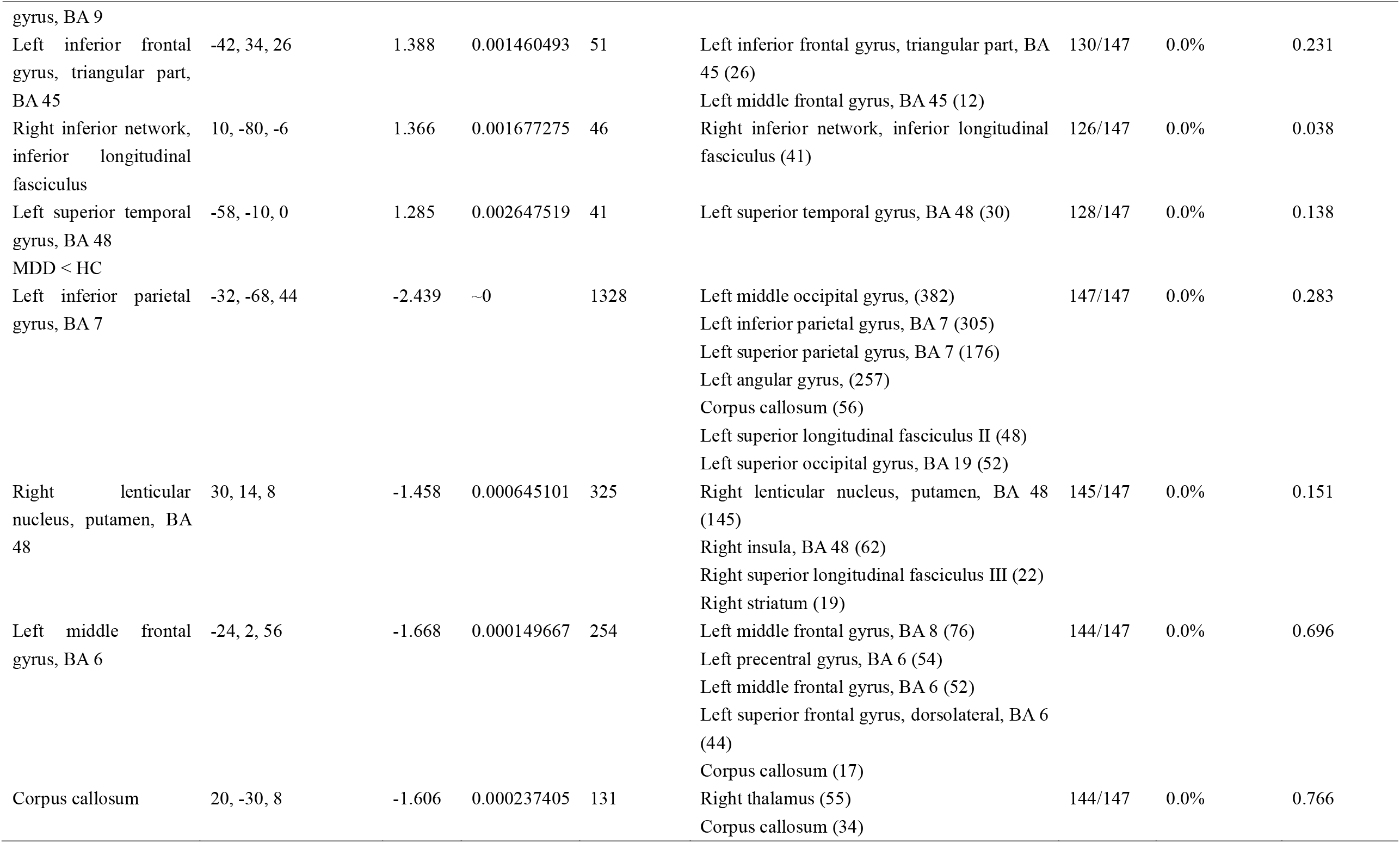
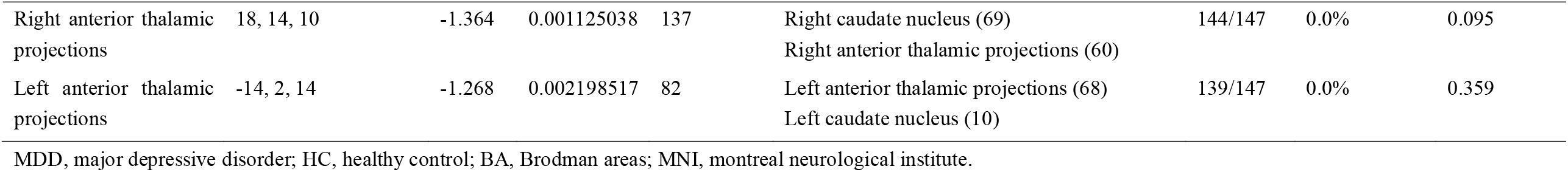
Clusters showing differences between MDD and HC.

#### 3.2.3. (rMDD vs. HCs) versus (MDD vs. HCs)

As illustrated in Fig. 1C and Table 3, the meta-analytic brain map illustrated both increased and decreased neural activation in rMDD patients relative to MDD patients. Relative to MDD patients, rMDD patients showed increased neural activation in the right MTG, right striatum, right caudate, left inferior parietal, and left median cingulate gyrus and decreased neural activation in the left cuneus cortex, right MTG, and left SFG (including the prefrontal cortex). These brain areas did not reveal obvious between-study heterogeneity. Egger’s tests also showed that there was no publication bias in each brain region (all *p* values > 0·05) (Table 3).

**Table 3.**
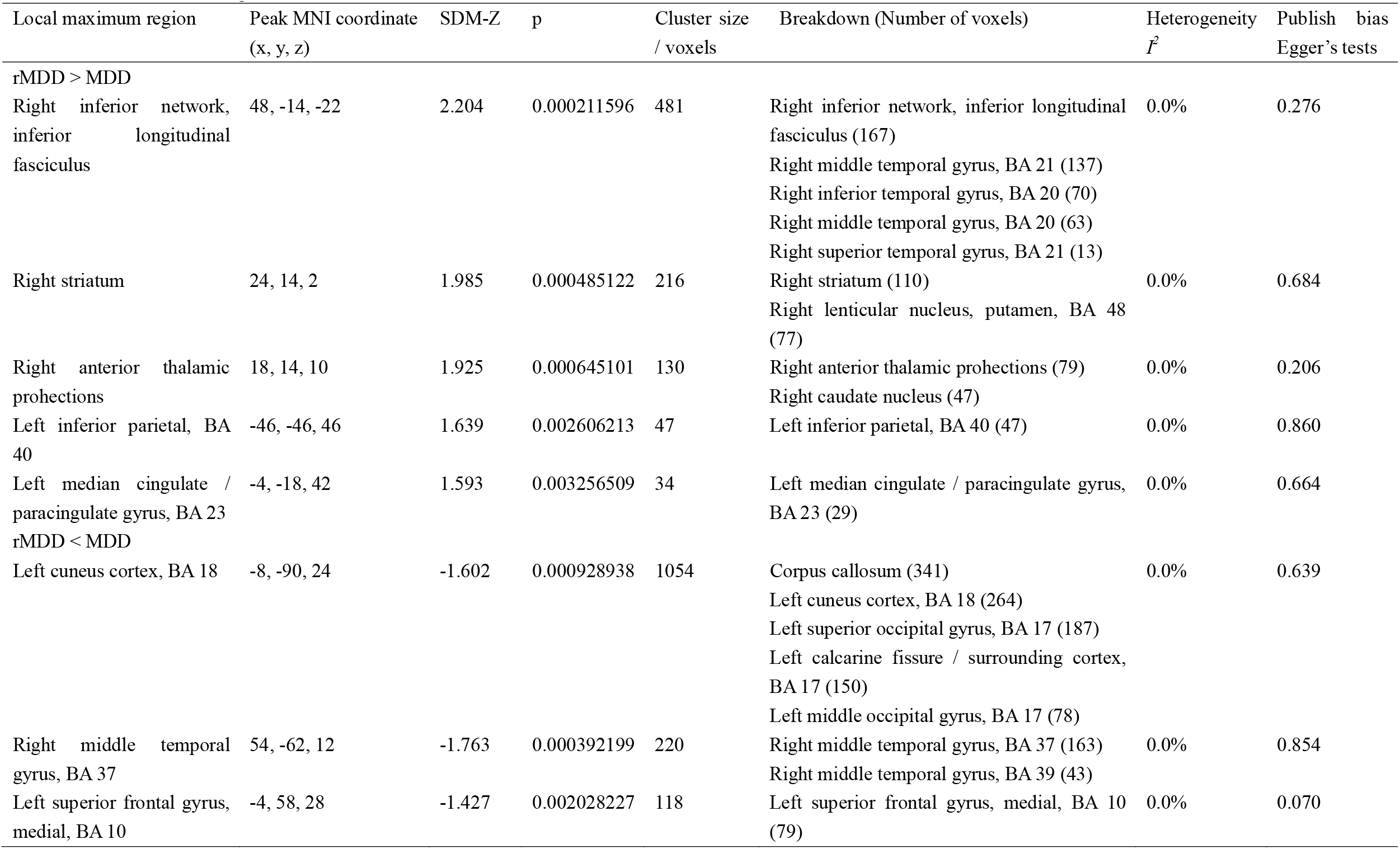
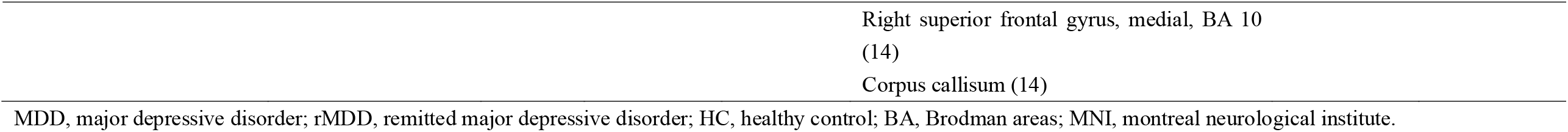
Clusters showing differences between (rMDD vs. HC) versus (MDD vs. HC).

#### 3.2.4. (rMDD vs. HCs) and (MDD vs. HCs) Conjunction

Conjunction analysis showed that compared with HCs, both rMDD and MDD patients had increased neural activation in the right temporal pole and right STG (Fig. 1D; right temporal pole: peak MNI = 54, 6, -14; *p* < 0.001; 288 voxels; right superior temporal gyrus: peak MNI = 50, -8, -12; *p* < 0.001; 282 voxels).

#### 3.2.5. Meta-regression Analysis

Meta-regression analyses revealed that percentage of females with MDD (available in all but six studies) was positively related with brain activation in the right lenticular nucleus/putamen (peak coordinates: x = 30, y = 14, z = 8; SDM + 1 per 0.004 point increase in the percentage of females, *p* = 0.0101) (Supplementary Fig. S2). We found no significant associations between the severity of depressive symptoms, mean age, or abnormal brain activity in patients with MDD. In rMDD patients, no significant associations were found between the number of depressive episodes, severity of depressive symptoms, average age, percentage of females, and brain regions with abnormal activation.

## 4. Discussion

To the best of our knowledge, this study is the first meta-analysis to investigate the common and distinct neural activation patterns of patients with MDD and patients with rMDD in task-related whole-brain fMRI studies. We found that MDD and rMDD patients shared increased neural activation in the right temporal pole and the right STG. Several brain regions, including the right MTG, left inferior parietal, prefrontal cortex, left SFG and the striatum differed significantly between MDD patients and rMDD patients.

### Comparison of rMDD Patients and HCs

Similar to previous task-related fMRI meta-analyses of rMDD patients (Wang et al., 2022), our results presented that compared with HCs, patients with rMDD showed significantly abnormal neural activaty in the left inferior parietal gyrus, left SFG, and right MTG, which play an important role in sensory perception, emotional control and cognitive functions, respectively. At present, the emotion regulation theory (van Kleef et al., 2022), and the cognitive model theory (Disner et al., 2011) are main hypotheses about the neuropathological mechanism of depression. Specifically, the left inferior parietal gyrus, SFG and MTG contribute to emotional control and cognitive function in MDD patients (Gong et al., 2020; Zheng et al., 2021). For rMDD individuals, stress may trigger a potential dysfunctional schema, in which the way of external and internal information is changed such that MDD patients prefer information with a certain emotional valence (or find such information aversive) in cognitive bias, thus promoting the recurrence of depressive mood states (Beck, 2008; van Kleef et al., 2022). Dysfunctional regulatory control and cognitive function may then prolong the experience of negative emotion, leading patients to spiral into a persistent depressed mood and finally a new depressive episode (Berking et al., 2014). Thus, disruptions in the processing and regulation of emotional information are important to understand how even mild adverse events may lead to subsequent MDD episodes. To a certain extent, our results of altered neural activation in task-related fMRI suggested that abnormalities in specific brain areas of rMDD patients can be used as biological indicators for predicting the recurrence of depression.

### Comparison of MDD Patients and HCs

Our results are consistent with previous task-related fMRI studies conducted in MDD patients, which reported that compared with those of HCs, the STG, insula, middle frontal gyrus (including the dorsolateral prefrontal cortex), IFG, and limbic system showed hypoactivation or hyperactivation in MDD patients (Diener et al., 2012; Fitzgerald et al., 2008; Müller et al., 2017). The insula is adjacent to the frontal lobe, temporal lobe, parietal lobe and limbic system, which are implicated in regulatory, affective, and disparate cognitive functions, including emotional responses, interoceptive awareness, and empathic processes (Gong et al., 2020). Moreover, many resting-state fMRI studies have shown altered functional connectivity in MDD patients involving the aforementioned abnormal neural activation, including the default mode network (DMN), salience network (SN), and central executive network (CEN) (Sha et al., 2019). The dynamic switching between the DMN and CEN involved in these brain regions facilitates access to cognitive resources, such as working memory and attention, when a salient event is detected. Thus, altered strength in the connection in these networks influences cognitive deficits in MDD patients (Gong et al., 2020). Previous task-based fMRI studies reported altered activation of the insula, dorsolateral prefrontal cortex, IFG, and limbic system during emotional processing tasks and executive functioning tasks in the brains of MDD patients (Miller et al., 2015a; Yang et al., 2022), which may partly explain the abnormalities in emotional and cognitive integration. In addition, altered activity in these brain areas occurs with a number of treatments for depression, including cognitive behavior therapy, deep brain stimulation, and medication, indicating a role for these regions in mediating the antidepressant response and remission more generally (McGrath et al., 2013). Moreover, the meta-analysis of current structural morphometric studies also revealed abnormal gray matter volume in these brain regions in MDD patients (Sha et al., 2019; Zhang et al., 2016). Therefore, structural and functional abnormalities in these areas may be critical neurobiological features of MDD.

### Differences between rMDD and MDD Patients

Consistent with previous task-related research, we observed different prefrontal cortex and SFG activation in MDD patients versus rMDD patients (Ming et al., 2017). Several resting-state fMRI studies also reported the aforementioned neural activity differences between MDD and rMDD patients (Yang et al., 2018; Young et al., 2014). Moreover, there is evidence of a negative association between the severity of depressive symptoms and reduced fMRI response of the prefrontal cortex and SFG in unaffected first-degree relatives of MDD and rMDD patients (Philippi et al., 2018; Watters et al., 2019). Based on the above results, it may be indicated that with the relief of depressive symptoms, abnormal activation of brain regions related to depressive symptoms also changes, which provides strong evidence for finding therapeutic targets for depression. It should be pointed out that the current research evidence from fMRI combined with artificial intelligence reported that depression is divided into different biotypes (Liang et al., 2020), and depression symptoms can actually be divided into different subgroups. However, to the best of our knowledge, there is no research exploring the relationship between the subgroups of depressive symptoms and abnormal neural activation, so it is necessary for future fMRI research to focus on the subgroups of depressive symptoms and explore their corresponding abnormal neural activation. This research is crucial for finding neurobiological targets for the precise treatment of depression.

Previous studies revealed that fMRI responses in the MTG, cingulate cortex, and prefrontal cortex during various tasks and resting-state valence significantly predicted posttreatment symptom improvement in individuals with MDD (Gao et al., 2022; Harris et al., 2022; Karim et al., 2018; Miller et al., 2015b). Our results also showed the different fMRI responses between rMDD and MDD patients in the MTG, cingulate cortex, and prefrontal cortex, which suggested that such differences may be due to neurological reactions after or during antidepressant treatment. However, most of the patients in the MDD group we included were receiving antidepressant treatment, while in the rMDD group, in only three studies were rMDD individuals receiving antidepressant treatment. Thus, an important direction of future research is to explore the differences in neural activation between first-episode and drug-naive MDD patients and rMDD patients who have stopped medication for a long period of time. The comparisons have important reference significance for finding the corresponding targets of specific antidepressant treatments, such as specific antidepressant drug therapy, physical therapy, and psychotherapy.

### Common Effects in Both rMDD and MDD Patients

The conjunction analysis suggested that neural activations of the right STG and right temporal pole were higher in both rMDD and MDD patients than in HCs, which may indicate a consistent inherent abnormal activity pattern across MDD states. Previous task-related fMRI studies implicating MDD and rMDD individuals also suggested abnormal neural activation of the right STG and right temporal pole (Dichter et al., 2012; Rai et al., 2021), which was further confirmed in our meta-analysis by comparing MDD and rMDD patients with HCs. The common disruptions in neural activation within both MDD and rMDD patients pose an interesting question for the study on the neuropathological mechanism of MDD: are the anomalously activated brain areas found in both conditions acquired alterations after MDD or intrinsic to MDD? If these abnormalities change after MDD acquisition and persist, they may represent objective neurophysiological markers for the diagnosis of depression. If these differences are intrinsic changes in the MDD population, this would infer that abnormal activatioy of related brain areas persists before the onset of MDD, during the course of MDD and after MDD remission, which may provide potential neurobiological markers for early screening of MDD. To the best of our knowledge, no longitudinally neuroimaging study has examined brain changes throughout the course of MDD (i.e., before, during and after remission); such studies are urgently needed to comprehensive understand the underlying mechanisms of MDD.

### Limitation

The current study was limited in some respects. The included studies were exclusive to those in adults; our findings may not be extrapolated to elderly individuals or children. Second, anomalous brain activation may gradually subside with the prolongation of the remission period (Ming et al., 2017), but only a few rMDD studies provided remission period information, which led us to fail to explore the impact of this aspect. Furthermore, the duration of MDD illness also affects neural activation (Zhou et al., 2017), but our meta-regression analysis did not find an effect of MDD course on neural activation. The remission period and the duration of illness are important issues that should be explored in future studies. Finally, the varieties in neroimaging sequence acquisition parameters, such as the number of coil channels, resolution, and field strength can lead to errors in statistical modelling. To mitigate these errors, it is essential to establish industry specifications akin to the Diagnostic and Statistical Manual of Mental Disorders (DSM) system to standardize methodology and ensure the results are applicable across different fields.

## 5. Conclusion

In conclusion, our meta-analysis shows that rMDD and MDD patients display a common pattern of aberrant task-related neural activation, which includes the right temporal pole and STG. Moreover, both rMDD and MDD patients exhibit distinct patterns of neural activation changes predominantly in the MTG, prefrontal cortex, and striatum. These findings seem to imply that the brain functions of rMDD and MDD patients have both similar and distinctive features and expands on a growing literature examining task-related fMRI in rMDD and MDD patients. Our results provide useful insights for understanding the potential neuropathology of brain dysfunction in MDD patients, developing more targeted and efficacious treatment and intervention strategies, and more importantly, providing potential neuroimaging targets for the early screening of MDD.

## Supporting information

Supplementary metarials

## Data Availability

All data produced in the present study are available upon reasonable request to the authors.

## ACKNOWLEDGEMENTS

This study was financially supported by the National Science Foundation of China (Grant No. 82171513); Institutional Research Fund from Sichuan University (No. 2022SCUH0026); Med-X Center for Informatics funding project (No. YGJC012); 1.3.5 project for disciplines of excellence, West China Hospital, Sichuan University (No. ZYJC21069) to Dr. H.Z.

