## Supplementary metarials for "Common And Distinct Patterns Of Task-Related Neural Activation Abnormalities In Patients With Remitted And Current Major Depressive Disorder: A Systematic Review And Coordinate-Based Meta-Analysis"

**Search formulas**

**Figure S1.** Flow chart of rMDD meta-analysis study selectionand MDD meta-analysis study selection

**Figure S2.** Meta-regression results

**Table S1.** Classifications of tasks according to the Research Domain Criteria (RDoC)

**Table S2.** Characteristics of included rMDD studies

**Table S3.** Characteristics of included MDD studies

**eReferences**

**Search formulas:**

rMDD:

Pubmed: ("MDD"[Title/Abstract] OR "major depressive disorder"[Title/Abstract] OR "unipolar depression"[Title/Abstract] OR "depressive disorder"[Title/Abstract]) AND ("remitted"[Title/Abstract] OR "remission" [Title/Abstract] OR "recovered" [Title/Abstract] OR "euthymic"[Title/Abstract]) AND ("functional MRI"[Title/Abstract] OR "fMRI" [Title/Abstract] OR "functional imaging" [Title/Abstract] OR "functional magnetic resonance imaging" [Title/Abstract] OR "neural" [Title/Abstract] OR "neuroimaging" [Title/Abstract])

Web of science: (AB=(major depressive disorder) OR AB=(mdd) OR AB=(unipolar depression) OR AB=(depressive disorder)) AND (AB=(fMRI) OR AB=(functional MRI) OR AB=(functional imaging) OR AB=(functional magnetic resonance imaging) OR AB=(neuroimaging)) AND (AB=(remitted) OR AB=(remission) OR AB=(recovered))

Embase: (‘mdd’:ab,ti OR ‘major depressive disorder’:ab,ti OR ‘unipolar depression’:ab,ti OR ‘depressive disorder’:ab,ti) AND (‘functional MRI’:ab,ti OR ‘fMRI’:ab,ti OR ‘functional imaging’:ab,ti OR ‘functional magnetic resonance imaging’:ab,ti OR ‘neuroimaging’:ab,ti) AND (‘remitted’:ab,ti OR ‘remission’:ab,ti OR ‘recovered’:ab,ti)

MDD:

Pubmed: ("MDD"[Title/Abstract] OR "major depressive disorder"[Title/Abstract] OR "unipolar depression"[Title/Abstract] OR "depressive disorder"[Title/Abstract]) AND ("functional MRI"[Title/Abstract] OR "fMRI" [Title/Abstract] OR "functional imaging" [Title/Abstract] OR "functional magnetic resonance imaging" [Title/Abstract] OR "neural" [Title/Abstract] OR "neuroimaging" [Title/Abstract])

Web of science: (AB=(major depressive disorder) OR AB=(mdd) OR AB=(unipolar depression) OR AB=(depressive disorder)) AND (AB=(fMRI) OR AB=(functional MRI) OR AB=(functional imaging) OR AB=(functional magnetic resonance imaging) OR AB=(neuroimaging))

Embase: (‘mdd’:ab,ti OR ‘major depressive disorder’:ab,ti OR ‘unipolar depression’:ab,ti OR ‘depressive disorder’:ab,ti) AND (‘functional MRI’:ab,ti OR ‘fMRI’:ab,ti OR ‘functional imaging’:ab,ti OR ‘functional magnetic resonance imaging’:ab,ti OR ‘neuroimaging’:ab,ti)

**eFigure 1.** Adapted Preferred Reporting Items for Systematic Reviews and Meta-Analyses (PRISMA) Flowchart Showing Study Inclusion. Flow chart of rMDD meta-analysis study selection (a) and MDD meta-analysis study selection (b).

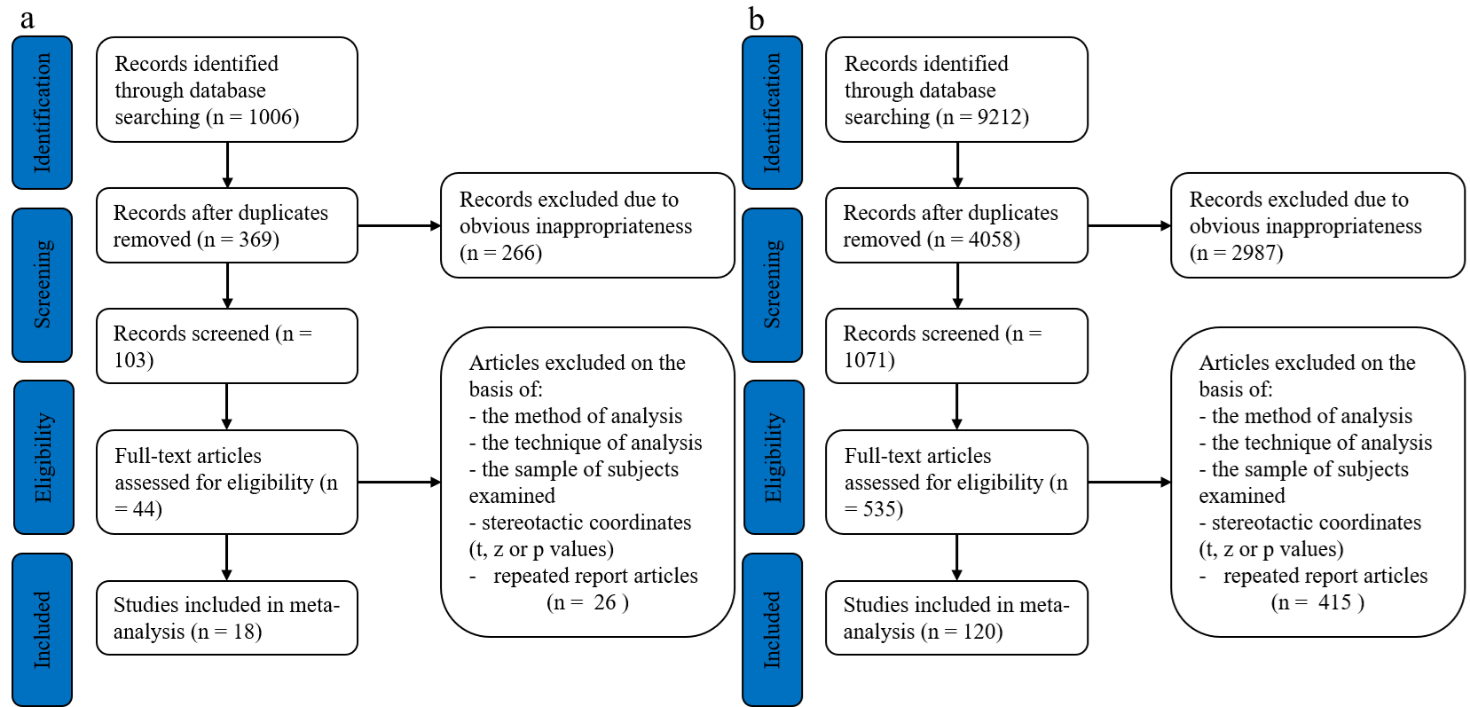

Abbreviation: MDD, major depressive disorder; rMDD, remitted major depressive disorder.

e**Figure 2.** Meta-regression results. Meta-regression results showing a positive association between female percentage and brain activity in right lenticular nucleus, putamen in major depressive disorder.

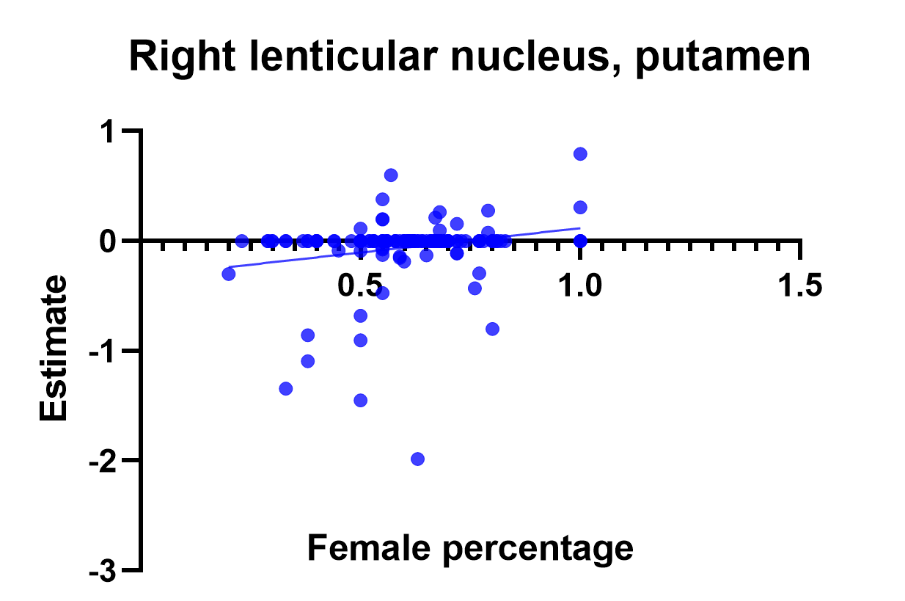

**eTable 1.** Classifications of tasks according to the Research Domain Criteria (RDoC).

| RDoC domain | RDoC Construct | Tasks |
| --- | --- | --- |
| Cognitive systems | Attention | Visual/Auditory oddball task, Attention shifting tasks, Continuous performance task |
|  | Memory | Episodic memory encoding/retrieval tasks, Semantic list learning tasks, Paired associates learning tasks |
|  | Language | Verbal fluency tasks, Semantic judgment tasks, Sentence Completion tasks |
|  | Cognitive control | Stroop task, Go/no-go task/ Stop task, Tower of London, Switching tasks |
|  | Working memory | N-back tasks, Sternberg task, Delayed-(non)-match-to-sample task |
|  | Cognitive unclassified | Implicit Learning tasks, Mental Arithmetic tasks |
| Positive valence systems | Approach/Motivation | Various tasks involving passive viewing of positively valenced scenes, words and facial expressions, Monetary incentive delay task (Reward expectation), Probabilistic reward task/Card-guessing task (Reward expectation) |
|  | Reward attainment | Monetary incentive delay task (Reward outcome); Reversal learning task (Reward outcome) |
| Negative valence systems | Acute threat | Fear conditioning, Symptom provocation |
|  | Potential threat | Various tasks involving passive viewing of negatively valenced scene, words and facial expressions, Monetary incentive delay task (Loss expectation) |
|  | Frustrative non-reward | Monetary incentive delay task (Punishment), Reversal learning task (Punishment), Probabilistic reward task (Punishment) |
| Social processes | Social communication | Various facial affect processing tasks |
|  | Perception and understanding of self | Various tasks involving processing of self-referential information |
|  | Perception and understanding of others | Theory of Mind tasks |
|  | Affiliation and attachment | Cyberball task |
| Cross domain tasks | Cross domain | Affective Go/NoGo, Emotional Stroop, Emotional identity task, Emotional Working memory task, Emotional Verbal Memory task, Emotional oddball task, Emotion attention shifting task, Emotion regulation task |

**eTable 2.** Characteristics of included rMDD studies

| Author/Year | PMID | Patients/Female | Controls/Female | Mean age ± SD Patients | Mean age ± SD Controls | Severity | Statistics | Medication status | Duration of illness (years) | Duration of remission (years) | Numbers of prior major depressive episodes | Software | Tal/MNI | Task | RDoC domain | RDoC Construct | Contrasts Used |
| --- | --- | --- | --- | --- | --- | --- | --- | --- | --- | --- | --- | --- | --- | --- | --- | --- | --- |
| Studies using experiments mapping the RDoC domains of negative valence systems and positive valence systems | | | | | | | | | | | | | | | | | |
| Admon et al., 2015 | 25483401 | 33/17 | 35/16 | 47.4 ± 1.8 | 45.7 ± 2.7 | HDRS-17 4.2 ± 4.2 | P < 0.005  uncorrected | No | 21.6 ± 9.3 | 7.3 ± 6.3 | 5.0 ± 2.3 | SPM8 | MNI | Negative valence during passive scene viewing | Negative valence | Potential Threat ("Anxiety") | Negative stimuli vs. baseline: rMDD > HC |
| Aust et al., 2013 | 24175181 | 14/9 | 14/8 | 55.1 ± 11.3 | 54.9 ± 11.8 | HDRS-17 3.8 ± 1.5 | P < 0.05  FDR Corrected | No | NA | NA | 3.9 ± 1.9 | SPM8 | MNI | Negative valence during passive scene viewing and auditory stimuli | Negative Valence | Potential threat | Unpleasant stimuli vs. neutral stimuli: rMDD > HC; rMDD < HC |
|  |  |  |  |  |  |  |  |  |  |  |  |  |  | Positive valence during passive scene viewing and auditory stimuli | Positive valence | Approach motivation | Pleasant stimuli vs. neutral stimuli: rMDD > HC; rMDD < HC |
| Schiller et al., 2013 | 23835103 | 19/15 | 19/12 | 23.6 ± 4.1 | 27.9 ± 6.3 | BDI 2.6 ± 4.9 | P < 0.005  uncorrected | No | NA | NA | 1.6 ± 0.9 | FSL4.1.8 | MNI | Monetary incentive delay task | Negative valence | Potential Threat | loss anticipation and outcomes vs. baseline: HC > rMDD |
| Smoski et al., 2013 | 23796796 | 18/14 | 19/12 | 24.8 ± 4.7 | 27.9 ± 6.3 | BDI 2.9 ± 5.0 | P < 0.005  uncorrected | No | NA | NA | 1.6 ± 0.9 | FSL4.1.8 | MNI | Negative valence during passive scene viewing | Negative Valence | Potential threat | Sad vs neutral: rMDD < HC |
| Dicther et al., 2012 | 22036801 | 19/15 | 19/12 | 23.6 ± 4.09 | 27.9 ± 6.3 | BDI 2.63 ± 4.91 | P < 0.005  uncorrected | No | NA | NA | 1.56 ± 0.86 | FSL4.1.8 | MNI | Monetary incentive delay task | Positive Valence | Approach Motivation | Reward anticipation (potential win vs non-potential win): rMDD > HC |
|  |  |  |  |  |  |  |  |  |  |  |  |  |  | Monetary incentive delay task | Positive valence | Initial Responsiveness to Reward Attainment | Reward outcome (wins vs non-wins): rMDD > HC; rMDD < HC |
| Rutgen et al., 2021 | 34049164 | 32/23 | 32/23 | 27.34 ± 1.35 | 27.41 ± 1.30 | HAMD 1.60 ± 0.48 | P < 0.05  FEW corrected | No | 6.1 ± 0.8 | NA | 1.8 ± 0.2 | SPM12 | MNI | Negative valence during passive scene viewing | Negative Valence | Potential threat | Painful vs neutral: rMDD < HC; rMDD > HC |
| Ming et al., 2017 | 28618857 | 33/17 | 36/18 | 21.67 ± 3.14 | 22.19 ± 1.60 | HRMD-17: 3.45 ± 2.73 | P < 0.05  FEW corrected | No | 0.91 ± 0.65 | 0.64 ± 0.52 | 1.73 ± 1.72 | SPM8 | MNI | Montreal Imaging Stress Task | Negative Valence | Potential threat | Stress vs Baseline: rMDD < HC; rMDD > HC |
| Albert et al., 2017 | 28012352 | 12/12 | 21/21 | 62.42 ± 5.79 | 60.64 ± 6.81 | BDI: 3.08 ± 3.26 | P < 0.001  corrected | No | 17.81 ± 10.85 | NA | 2.1 ± 0.95 | SPM8 | MNI | Negative valence during passive scene viewing | Negative Valence | Potential threat | Negative images vs Neutral images: rMDD > HC |
| Jenkins et al., 2018 | 28826089 | 48/32 | 40/25 | 21.36/- | 20.88/- | HAMD: 1.84 | P < 0.05  FEW corrected | No | NA | NA | NA | SPM8 | MNI | Negative valence during passive scene viewing | Negative Valence | Potential threat | Sad, fear or anger stimuli vs. baseline: rMDD > HC; rMDD < HC |
|  |  |  |  |  |  |  |  |  |  |  |  |  |  | Positive valence during passive scene viewing | Positive valence | Approach motivation | Happy vs. baseline: rMDD < HC |
| Studies using experiments mapping the RDoC social processes | | | | | | | | | | | | | | | | | |
| Nejad et al., 2019 | 30641339 | 25/21 | 29/25 | 38.8 ± 12.0 | 41.4 ± 12.8 | BDI ≤10 or MADRS≤7 | P < 0.05  FEW corrected | No | 11.1/- | 0.93 ± 0.59 | 3.2 ± 2.4 | SPM8 | MNI | Processing of self-referential information | Social Processes | Perception and Understanding of Self | Self-perception vs Baseline: rMDD < HC |
| Norbury et al., 2010 | 19627640 | 16/9 | 21/10 | 36.2 ± 13.9 | 32.3 ± 12.9 | BDI: 3.5 ±3.7 | P < 0.05 corrected | No | NA | NA | NA | FSL | MNI | Unspecified valence during facial affect processing | Social process | Social communication | Fearful vs. happy facial expressions: rMDD > HCs |
| Studies using experiments mapping the RDoC cognitive systems domain | | | | | | | | | | | | | | | | | |
| Dai et al., 2018 | 29575563 | 27/17 | 33/21 | 49.56 ± 10.77 | 48.85 ± 12.86 | HDRS: 5.11 ± 1.91 | P < 0.05  FEW corrected | No | 9.76 ± 5.16 | NA | 2.56 ± 1.39 | SPM8 | MNI | Inhibition of return | Cognitive Systems | Cognitive Control | Invalid cue vs. valid cue:  rMDD > HC |
| Jenkins et al., 2016 | 26714574 | 32/21 | 25/14 | 21.53 ± 1.57 | 21.12 ± 1.83 | HAMD 1.77 ± 1.89 | P < 0.05  corrected | No | NA | NA | 2/- | SPM8 | MNI | Switching tasks | Cognitive Systems | Cognitive Control | Facial emotion perception vs Baseline: rMDD > HC |
| Young et al., 2015 | 25919972 | 20/12 | 20/13 | 30 ± 12 | 29 ± 9 | HDRS: 3.40 ± 4.17 | P < 0.05  corrected | No | NA | NA | NA | AFNI | Tal | Episodic memory retrieval task | Cognitive Systems | Memory | Specific memory vs. Example Generation: HC > rMDD |
|  |  |  |  |  |  |  |  |  |  |  |  |  |  | Episodic memory retrieval task | Cognitive Systems | Memory | Positive Specific memory vs. Positive Example Generation: HC > rMDD |
|  |  |  |  |  |  |  |  |  |  |  |  |  |  | Episodic memory retrieval task | Cognitive Systems | Memory | Negative Specific memory vs. Negative Example Generation: rMDD > HC; HC > rMDD |
| Breukelaar et al., 2020 | 31604366 | 25/13 | 25/16 | 34.82 ± 11.04 | 34.86 ± 12.63 | HDRS: 6.28 ± 4.23 | P < 0.05  FEW corrected | Yes | NA | NA | 11.88 ± 15.57 | SPM8 | MNI | Working memory task | Cognitive systems | Working memory | Working memory task vs. baseline: MDD > HCs |
| Schöning et al., 2009 | 19086021 | 28/16 | 28/16 | 34.18 ± 10.62 | 33.42 ± 9.62 | HAMD: 3.64 ± 2.63 | P < 0.0005  uncorrected | Yes | NA | NA | 2.54 ± 1.75 | SPM5 | MNI | N-back task | Cognitive system | Working memory | 2-back vs. 0 and 1-back: MDD > HCs |
| Study including cross-domain experiments | | | | | | | | | | | | | | | | | |
| Smoski et al., 2015 | 25617820 | 18/14 | 19/12 | 24.8 ± 4.7 | 27.9 ± 6.3 | BDI 2.9 ± 5.0 | P < 0.005  uncorrected | No | NA | NA | 1.6 ± 0.9 | FSL4.1.8 | MNI | Emotion regulation task | Cognitive affective interaction | Cognitive affective interaction | Acceptance vs reappraising: rMDD < HC |
|  |  |  |  |  |  |  |  |  |  |  |  |  |  | Emotion regulation task | Cognitive affective interaction | Cognitive affective interaction | Acceptance vs Viewing: rMDD < HC |
| Rai et al., 2021 | 33369067 | 39/20 | 41/21 | 36.7 ± 12.2 | 36.7 ± 13.8 | HDRS: 4.62 ± 3.82 | P < 0.05  FEW corrected | Yes | NA | NA | 5.16 ± 6.4 | SPM12 | MNI | Emotion regulation task | Cognitive affective interaction | Cognitive affective interaction | Watch or think vs. netural or watch: rMDD > HC |

rMDD, remitted major depressive disorder; HC, healthy control; MNI, montreal neurological institute; BDI, Beck depression inventory; MADR, Montgomery–Asberg Depression Rating Scale; HAMD, Hamilton depression scale; SD, standard deviation; RDoC, Research domain criteria

**Table S3.** Characteristics of included MDD studies

| Author/Year | PMID | Patients/Female | Controls/Female | Mean age ± SD Patients | Mean age ± SD Controls | Severity mean ± SD | Duration of illness (years) | Numbers of major depressive episodes | Statistics | Medication status | Software | Tal/MNI | Task | RDoC domain | RDoC Construct | Contrasts Used |
| --- | --- | --- | --- | --- | --- | --- | --- | --- | --- | --- | --- | --- | --- | --- | --- | --- |
| Studies using experiments mapping the RDoC domains of negative valence systems and positive valence systems | | | | | | | | | | | | | | | | |
| Abler et al., 2007 | 17010993 | 12/12 | 12/12 | 41.2/- | 40.7/- | HAMD: 18.5 (median) | Unknown | Unknown | P < 0.05  corrected | Yes | SPM2 | MNI | Negative valence during passive scene viewing | Negative valence | Potential threat | Presentation negative vs. neutral stimuli, Expectation negative vs. neutral stimuli: MDD > HCs |
|  |  |  |  |  |  |  |  |  |  |  |  |  | Positive valence during passive scene viewing | Positive valence | Approach motivation | Presentation positive vs. neutral stimuli: MDD > HCs |
| Arrondo et al., 2015 | 26379600 | 24/7 | 21/4 | 33.8 ± 9.15 | 34.33 ± 10.11 | BDI: 32/- | Unknown | Unknown | P < 0.05  uncorrected | Yes | FSL | MNI | Monetary incentive delay task | Positive valence | Approach motivation | Reward anticipation vs. anticipation of a neutral outcome: MDD < HCs |
| Wang et al., 2022 | 35079855 | 23/15 | 30/18 | 28.61± 9.19 | 26.87 ± 4.97 | HAMD: 14.52 ± 8.74 | 2.52 ± 2.99 | Unknown | P < 0.05  FEW corrected | Yes | SPM | MNI | Monetary incentive delay task (reward outcome) | Positive valence | Reward attainment | Reward magnitude vs. baseline:  MDD > HCs |
| Li et al., 2022 | 34888973 | 37/25 | 37/19 | 31.21 ± 12.17 | 36.57 ± 16.28 | HAMD: 20.11 ± 6.90 | 2.99 ± 2.77 | Unknown | P < 0.05  FDR corrected | Yes | SPM12 | MNI | Negative valence during passive scene viewing | Negative Valence | Potential threat | Emotionally incongruent vs. emotionally congruent: MDD < HCs |
|  |  |  |  |  |  |  |  |  |  |  |  |  | Positive valence during passive scene viewing | Positive valence | Approach motivation | Emotionally incongruent vs. emotionally congruent: MDD > HCs; MDD < HCs |
| Trettin et al., 2022 | 34728281 | 36/21 | 30/20 | 40.71 ± 12.72 | 39.47 ± 13.19 | BDI: 18.50 ± 1.43 | 11.92 ± 12.25 | 12.40 ±3.35 | P < 0.01  Cluster-level corrected | Yes | brainvoyager | Tal | Negative valence during passive scene viewing | Negative Valence | Potential threat | Negative stimuli vs. baseline: MDD < HCs |
| Reinen et al., 2021 | 34517334 | 24/12 | 24/12 | 26.58 ± 6.40 | 26.89 ± 5.50 | HAMD-17: 20.08 ± 2.59 | 8.81/- | Unknown | P < 0.05  FEW corrected | No | SPM8 | MNI | Reward attainment | Positive valence | Reinforcement learning task (reward outcome) | Gain vs. baseline (overall prediction error): MDD < HCs |
|  |  |  |  |  |  |  |  |  |  |  |  |  | Frustrative Non-reward | Negative Valence | Probabilistic reward task (Punishment) | Loss vs. baseline (overall prediction error): MDD < HCs; MDD > HCs |
| Rutgen et al., 2021 | 34049164 | 29/21 | 32/23 | 29.62 ± 1.76 | 27.41 ± 1.30 | HAMD: 25.90 ± 1.19 | 9.6 ± 1.7 | 3.4 ± 0.4 | P < 0.05  FEW corrected | No | SPM12 | MNI | Negative valence during passive scene viewing | Negative Valence | Potential threat | Painful vs. neutral: MDD < HCs; MDD > HCs |
| Rupprechter et al., 2021 | 31907081 | 18/15 | 16/10 | 18-33 | 17-41 | BDI: 25.9 ± 12.9 | Unknown | Unknown | P < 0.01  Monte Carlo method corrected | No | SPM12 | Unknown | Probabilistic reward task | Positive valence | Reward attainment | Reward expectancy vs. baseline: MDD < HCs |
| Gao et al., 2021 | 33418366 | 55/35 | 44/17 | 21.16 ± 2.02 | 20.02 ± 1.19 | BDI: 29.44 ± 8.23 | 1.8/- | 1.00 ±1.22 | P < 0.05  FEW corrected | Yes | SPM12 | MNI | Probabilistic reward task | Positive valence | Reward attainment | Win outcomes vs. baseline: MDD < HCs |
| Tak et al., 2021 | 33403894 | 34/34 | 28/28 | 24.5 ± 2.8 | 24.4 ± 2.6 | BDI: 33.7 ± 7.1 | Unknown | Unknown | P < 0.005  uncorrected | No | SPM12 | MNI | Negative valence during passive scene viewing | Negative Valence | Potential threat | Negative vs. neutral images: MDD > HCs; MDD < HCs |
| Mel'nikov et al., 2018 | 30353344 | 21/- | 21/- | Unknown | Unknown | Unknown | Unknown | Unknown | P < 0.05  FEW corrected | No | SPM8 | MNI | Negative valence during passive scene viewing | Negative valence | Potential threat | Sad images vs. netural: MDD < HCs |
| Strigo et al., 2013 | 23481626 | 31/15 | 22/11 | 27.6 ± 7.8 | 26.8 ± 8.7 | BDI: 25.5 ± 8.4 | Unknown | 2 ± 1 | P < 0.05 corrected | No | Other | Tal | Pain-anticipation task | Negative valence | Potential threat | High vs. low pain anticipation: MDD < HCs; MDD > HCs |
| Deldonno et al., 2019 | 31170437 | 23/16 | 27/23 | 25.09 ± 3.32 | 29.15 ± 9.0 | HAMD: 18.56 ± 3.24 | Unknown | 4.25 ±2.53 | P < 0.01  uncorrected | No | SPM8 | MNI | Monetary incentive delay task | Positive valence | Approach/Motivation | Win vs. neutral: MDD < HCs |
| Ming et al., 2017 | 28618857 | 36/19 | 36/18 | 22.81 ± 4.25 | 22.19 ± 1.60 | HAMD-17: 22.56 ± 5.64 | 0.61± 0.44 | Unknown | P < 0.05  FEW corrected | No | SPM8 | MNI | Montreal Imaging Stress Task | Negative Valence | Potential threat | Stress vs. Baseline: MDD < HCs; MDD > HCs |
| Engelmann et al., 2017 | 28587695 | 19/10 | 23/14 | 37.6 ± 11.0 | 33.7 ± 11.6 | HAMD: 22.84 ± 4.17 | Unknown | Unknown | P < 0.05  FEW corrected | No | SPM8 | MNI | Probabilistic reward task | Negative Valence | Frustrative non-reward | Loss related recoding vs. baseline: MDD > HCs |
| Young et al., 2017 | 28446254 | 40/20 | 40/- | 35.8/- | 18-55 | HDRS: 19.2/- | Unknown | Unknown | P < 0.05  corrected | No | AFNI | Tal | Positive valence during passive scene viewing | Positive valence | Approach/Motivation | Positive specific memories vs. positive example generation: MDD < HCs; MDD > HCs |
|  |  |  |  |  |  |  |  |  |  |  |  |  | Negative valence during passive scene viewing | Negative Valence | Potential threat | Negative specific memories vs. negative example generation: MDD > HCs |
| Lawson et al., 2017 | 27240528 | 25/10 | 15/11 | 27.76 ± 9.01 | 27.44 ± 8.75 | HAMD: 19.08 ± 3.75 | Unknown | 2.88 ± 1.01 | P < 0.01  uncorrected | No | SPM8 | MNI | Positive valence during passive scene viewing | Positive valence | Approach/Motivation | Win vs. neutral: MDD < HCs; MDD > HCs |
|  |  |  |  |  |  |  |  |  |  |  |  |  | Negative valence during passive scene viewing | Negative Valence | Potential threat | Lose vs. neutral: MDD < HCs; MDD > HCs |
| Bürger et al., 2017  n/p | 28205606 | 36/22 | 36/19 | 40.72 ± 11.58 | 41.33 ± 6.05 | HAMD: 23.61 ± 3.17 | 11.07 ± 8.95 | 4.61 ±3.66 | P < 0.05  FEW corrected | Yes | SPM8 | MNI | Positive valence during passive scene viewing | Positive valence | Approach/Motivation | Happy vs. neutral: MDD < HCs |
|  |  |  |  |  |  |  |  |  |  |  |  |  | Negative valence during passive scene viewing | Negative Valence | Potential threat | Fearful vs. neutral: MDD < HCs |
| Liu et al., 2017 | 28575424 | 21/12 | 17/10 | 30.7 ± 8.9 | 29.3 ± 5.2 | HDRS: 24.05 ± 4.15 | 2.90±2.65 | 1.42±0.59 | P < 0.05  FEW corrected | No | SPM8 | MNI | Negative valence during passive scene viewing | Negative Valence | Potential threat | Punishment vs. neutral: MDD > HCs |
| Chandrasekhar Pammi et al., 2015 | 25923684 | 10/2 | 10/4 | 31.9 ± 7.5 | 27.5 ± 2.4 | Unknown | Unknown | Unknown | P < 0.001  uncorrected | Unknown | SPM8 | MNI | Probabilistic reward task | Positive valence | Initial responsiveness to reward attainment | Gain vs. loss: MDD < HCs; MDD > HCs |
| Chase et al., 2013 | 24148027 | 40/31 | 40/25 | 31.04 ± 8.04 | 33.09 ± 6.23 | HDRS: 26.63 ± 5.70 | 12.92 ± 7.25 | Unknown | P < 0.005  uncorrected | Yes | SPM8 | MNI | Card-guessing task | Positive valence | Approach motivation | Reward expectancy vs. baseline: MDD < HCs; MDD > HCs |
| Demenescu et al., 2011 | 21557888 | 59/39 | 56/34 | 36.24 ± 10.79 | 39.75 ± 9.67 | MADRS: 11.16 ± 8.66 | Unknown | Unknown | P < 0.05  FDR corrected | Yes | SPM5 | MNI | Positive valence during facial affect processing | Positive valence | Approach motivation | Happy vs. scram bled: MDD > HCs |
|  |  |  |  |  |  |  |  |  |  |  |  |  | Unspecified valence during facial affect processing | Social process | Social communication | Neutral vs. scrambled: MDD > HCs |
| Derntl et al., 2011 | 21777105 | 15/9 | 15/9 | 34.1 ± 11.95 | 32.9 ± 10.93 | HAMD: 19.9 ± 7.30 | 4.7 ±7.12 | Unknown | P < 0.05  corrected | Yes | SPM5 | MNI | Positive valence during passive scene viewing | Positive valence | Approach motivation | Happy vs. neutral: MDD < HCs; MDD > HCs |
|  |  |  |  |  |  |  |  |  |  |  |  |  | Negative valence during passive scene viewing | Negative valence | Potential threat | Anger vs. neutral: MDD < HCs |
| Frodl et al., 2009 | 17965984 | 12/7 | 12/7 | 43.3 ± 11.2 | 41.3 ± 11.7 | HAMD: 17.5 ± 4.4 | Unknown | Unknown | P < 0.001  uncorrected | Yes | SPM2 | MNI | Negative valence during passive scene viewing | Negative valence | Potential threat | Emotional processing vs. match forms: MDD > HCs |
| Fu et al., 2007  n/p | 17403973 | 19/13 | 19/11 | 43.2 ± 8.8 | 42.8 ± 6.7 | HAMD: 21.1 ± 2.3 | Unknown | Unknown | P < 0.005  corrected | No | Unknown | Tal | Positive valence during passive scene viewing | Positive valence | Approach motivation | Happy Faces vs. baseline: MDD < HCs |
| Fu et al., 2008  n/p | 18550030 | 16/13 | 16/13 | 39.2 ± 9.3 | 40.0 ± 9.4 | HAMD: 20.9 ± 1.9 | Unknown | Unknown | P < 0.005  corrected | No | Unknown | Tal | Negative valence during passive scene viewing | Negative valence | Potential threat | Sad facial affect vs. baseline: MDD < HCs; MDD > HCs |
| Gotlib et al., 2005 | 16237317 | 18/13 | 18/13 | 35.2/- | 30.8/- | BDI: 24.6 ± 8.3 | Unknown | Unknown | P < 0.001  uncorrected | Yes | SPM99 | Tal | Positive valence during passive scene viewing | Positive valence | Approach motivation | Happy vs. neutral faces: MDD < HCs; MDD > HCs |
|  |  |  |  |  |  |  |  |  |  |  |  |  | Negative valence during passive scene viewing | Negative valence | Potential threat | Sad vs. neutral faces: MDD < HCs; MDD > HCs |
| Greening et al., 2013 | 23769293 | 18/12 | 18/12 | 26.61 ± 11.70 | 27.89 ± 11.26 | BDI: 25.53 ± 10.4 | Unknown | Unknown | P < 0.05  FEW corrected | Yes | AFNI | MNI | Negative valence during passive scene viewing | Negative valence | Potential threat | Response to fearful task-irrelevant distracters vs. baseline: MDD < HCs |
| Johnston et al., 2015 | 26133661 | 19/15 | 21/15 | 50.79 ± 10.6 | 46.14 ± 13.97 | HAMD: 16.00 ± 5.72 | Unknown | Unknown | P < 0.01  FEW corrected | Yes | SPM8 | MNI | Probabilistic reward task | Positive valence | Initial responsiveness to reward attainment | Win vs. control: MDD < HCs; MDD > HCs |
|  |  |  |  |  |  |  |  |  |  |  |  |  | Probabilistic reward task | Negative valence | Frustrative nonreward | Loss vs. control: MDD < HCs; MDD > HCs |
| Keedwell et al., 2005 | 15993859 | 12/8 | 12/8 | 43 ± 9.8 | 36 ± 14.6 | BDI: 33.5 ± 11.2 | Unknown | Unknown | P < 0.01  uncorrected | Yes | Other | Tal | Positive valence during facial affect processing | Positive valence | Approach motivation | Happy mood vs. neutral conditions: MDD < HCs; MDD > HCs |
|  |  |  |  |  |  |  |  |  |  |  |  |  | Negative valence during facial affect processing | Negative valence | Potential threat | Sad mood vs. neutral conditions: MDD < HCs; MDD > HCs |
| Zhong et al., 2012 | 22398297 | 27/16 | 25/14 | 20.37 ± 1.86 | 20.96 ± 1.54 | CES-D: 25.1 ± 15.42 | Unknown | Unknown | P < 0.001  uncorrected | No | SPM8 | MNI | Negative valence during facial affect processing | Negative valence | Potential threat | Matching fearful and angry faces vs. matching forms: MDD < HCs; MDD > HCs |
| Murrough et al., 2015 | 25689570 | 18/8 | 20/9 | 38.1 ± 13.8 | 35.0 ± 8.9 | MADRS: 29.9 ± 6.8 | 24.2 ± 15.7 | 2.4 ± 1.7 | P < 0.05  FEW corrected | Yes | SPM8 | MNI | Positive valence during facial affect processing | Positive valence | Approach motivation | Happy vs. neutral: MDD < HCs |
| Pizzagalli et al., 2009 | 19411368 | 30/15 | 31/13 | 43.17 ± 12.98 | 38.80 ± 14.48 | HAMD: 17.97 ± 4.9 | Unknown | 3.69 ± 2.64 | P < 0.005 corrected | No | FreeSurfer | MNI | Monetary incentive delay task | Positive valence | Approach motivation | Reward cue vs. no incentive cue: MDD < HCs; MDD > HCs |
|  |  |  |  |  |  |  |  |  |  |  |  |  | Monetary incentive delay task | Positive valence | Initial responsiveness to reward attainment | Gain vs. no-change feedback: MDD < HCs; MDD > HCs |
|  |  |  |  |  |  |  |  |  |  |  |  |  | Monetary incentive delay task | Negative valence | Potential threat | Loss cue vs. no incentive cue: MDD < HCs; MDD > HCs |
|  |  |  |  |  |  |  |  |  |  |  |  |  | Monetary incentive delay task | Negative valence | Frustrative nonreward | Penalty vs. no-change feedback: MDD < HCs; MDD > HCs |
| Remijnse et al., 2009 | 19171077 | 20/8 | 27/19 | 35/- | 32/- | HAMD: 19.1 ± 4.1 | Unknown | Unknown | P < 0.001  uncorrected | No | SPM2 | MNI | Reversal learning task | Positive valence | Initial responsiveness to reward attainment | Reward vs. baseline: MDD > HCs |
|  |  |  |  |  |  |  |  |  |  |  |  |  | Reversal learning task | Negative valence | Frustrative nonreward | Punishment vs. baseline: MDD < HCs; MDD > HCs |
| Rizvi et al., 2013 | 23948629 | 21/14 | 18/12 | 38.9 ± 11.4 | 36.2 ± 10.3 | HAMD: 21.8 ± 1.5 | Unknown | Unknown | P < 0.05  uncorrected | Yes | FSL | MNI | Positive valence during passive scene viewing | Positive valence | Approach motivation | Positive images vs. neutral: MDD > HCs |
|  |  |  |  |  |  |  |  |  |  |  |  |  | Negative valence during passive scene viewing | Negative valence | Potential threat | Negative images vs. neutral: MDD > HCs |
| Robinson et al., 2012 | 22420038 | 13/5 | 14/6 | 36 ± 11 | 31 ± 6 | HAMD: 20 ± 7 | Unknown | Unknown | P < 0.001  uncorrected | No | SPM8 | MNI | Reversal learning task | Positive valence | Initial responsiveness to reward attainment | Unexpected reward vs. baseline: MDD < HCs |
| Scheuerecker et al., 2010 | 20569645 | 13/3 | 15/5 | 37.9 ± 10.1 | 35.5 ± 10.9 | HAMD: 20.5 ± 4.7 | 4.36 ± 5.96 | 1.45 ± 0.68 | P < 0.001  uncorrected | No | SPM5 | MNI | Negative valence during facial affect processing | Negative valence | Potential threat | Sad or angry faces vs. shapes: MDD > HCs |
| Segarra et al., 2016 | 26708106 | 24/7 | 21/4 | 33.08 ± 9.15 | 34.33 ± 10.11 | BDI: 32.62 ± 7.06 | Unknown | Unknown | P < 0.05 FWE  corrected | Yes | FSL | MNI | Probabilistic reward task | Positive valence | Initial responsiveness to reward attainment | Win outcomes vs full miss outcomes: MDD < HCs |
| Smoski et al., 2009 | 19261334 | 14/7 | 15/9 | 34.8 ± 14.3 | 30.8 ± 9.7 | HAMD: 23.5/- | Unknown | Unknown | Z > 2.6 corrected | No | FSL | Tal | Probabilistic reward task | Positive valence | Approach motivation | Anticipation phase (Money vs. control trials): MDD < HCs; MDD > HCs |
|  |  |  |  |  |  |  |  |  |  |  |  |  | Probabilistic reward task | Positive valence | Initial responsiveness to reward attainment | Feedback phase (Winning vs. control trials): MDD < HCs; MDD > HCs |
|  |  |  |  |  |  |  |  |  |  |  |  |  | Probabilistic reward task | Negative valence | Frustrative nonreward | Feedback phase (Non-win vs. control trials): MDD < HCs; MDD > HCs |
| Smoski et al., 2011 | 22079658 | 9/- | 13/- | 34.4 ± 15.1 | 26.2 ± 6.3 | BDI: 16.7 ± 4.9 | Unknown | Unknown | Z > 2.58 corrected | Yes | FSL | Tal | Monetary incentive delay task | Positive valence | Approach motivation | Anticipation phase, money and Images (potential win vs. non-potential win): MDD < HCs; MDD > HCs |
|  |  |  |  |  |  |  |  |  |  |  |  |  | Monetary incentive delay task | Positive valence | Initial responsiveness to reward attainment | Reward outcome, money and Images (win vs. non-win): MDD < HCs; MDD > HCs |
| Surguladze et al., 2005 | 15691520 | 16/6 | 14/6 | 42.3 ± 8.4 | 35.1 ± 13.2 | BDI: 31.1 ± 10.8 | 7.5 ± 5.1 | Unknown | P < 0.05  corrected | Yes | Other | Tal | Positive valence during facial affect processing | Positive valence | Approach motivation | Response to increasing intensities to happy facial expressions vs. baseline: MDD < HCs |
|  |  |  |  |  |  |  |  |  |  |  |  |  | Negative valence during facial affect processing | Negative valence | Potential threat | Response to increasing intensities to sad facial expressions vs. baseline: MDD > HCs |
| Surguladze et al., 2010 | 20307892 | 9/4 | 9/4 | 42.8 ± 7.2 | 39.7 ± 14.6 | HAMD: 17.7 ± 5.5 | 8.0 ± 5.1 | Unknown | P < 0.05  corrected | Yes | Other | Tal | Negative valence during facial affect processing | Negative valence | Potential threat | Disgust and fear vs. neutral faces: MDD < HCs; MDD > HCs |
| Townsend et al., 2010 | 20708906 | 15/9 | 15/9 | 45.6 ± 11.2 | 44.8 ± 11.7 | HAMD: 20.1 ± 4.9 | 14.7 ± 13.3 | 3  (median) | P < 0.05  corrected | No | FSL | Tal | Negative valence during facial affect processing | Negative valence | Potential threat | Match fearful or sad faces vs. match forms: MDD < HCs |
| Victor et al., 2010 | 23056309 | 22/12 | 25/15 | 31 ± 7.8 | 29 ± 6.7 | HAMD: 24 ± 6.3 | Unknown | 3.2 ± 2.0 | P < 0.001  uncorrected | Yes | SPM5 | Tal | Positive valence during facial affect processing | Positive valence | Approach motivation | Masked-happy faces vs. masked-neutral faces: MDD < HCs; MDD > HCs |
|  |  |  |  |  |  |  |  |  |  |  |  |  | Negative valence during facial affect processing | Negative valence | Potential threat | Masked-sad vs. masked-neutral: MDD < HCs; MDD > HCs |
| Wang et al., 2012 | 22954751 | 18/11 | 18/11 | 31.61 ± 7.88 | 31.67 ± 6.82 | Unknown | Unknown | Unknown | P < 0.001  uncorrected | No | SPM5 | MNI | Positive valence during passive scene viewing | Positive valence | Approach motivation | Positive stimuli vs. baseline: MDD < HCs |
|  |  |  |  |  |  |  |  |  |  |  |  |  | Negative valence during passive scene viewing | Negative valence | Potential threat | Negative stimuli vs. baseline: MDD > HCs |
| Yang et al., 2016 | 26192817 | 25/13 | 25/15 | 28.96 ± 7.00 | 28.36 ± 7.87 | HAMD: 27.58 ± 4.62 | Unknown | 1/- | P < 0.05 AlphaSim  corrected | No | SPM8 | MNI | Probabilistic reward task | Positive valence | Approach motivation | Prob8 vs. rest: MDD < HCs |
|  |  |  |  |  |  |  |  |  |  |  |  |  | Probabilistic reward task | Positive valence | Initial responsiveness to reward attainment | High reward vs. rest: MDD < HCs |
| Zhong et al., 2011 | 21878364 | 29/16 | 31/16 | 20.45 ± 1.82 | 20.84 ± 1.49 | CES-D: 34.86 ± 5.41 | Unknown | 1/- | P < 0.05 FWE  corrected | No | SPM8 | MNI | Negative valence during facial affect processing | Negative valence | Potential threat | Matching fearful or angry faces vs. matching forms: MDD < HCs; MDD > HCs |
| Goodon et al., 2019 | 30890968 | 13/10 | 14/8 | 30.6 ± 10.0 | 29.8 ± 9.6 | - | Unknown | 9.23 ± 13.22 | P < 0.05  corrected | Yes | FSL | MNI | Positive valence during passive scene viewing | Positive valence | Approach motivation | Happy vs. baseline: MDD < HCs |
| Nagy et al., 2021 | 34646916 | 21/14 | 21/14 | 32.52 ± 9.55 | 33.24 ± 8.37 | BDI: 23.10 ± 5.66 | 4/- | 2 /- | P < 0.05  FEW corrected | No | FSL | MNI | Negative valence during passive scene viewing | Negative valence | Potential threat | Images portraying negative emotions vs. baseline: MDD < HCs; MDD > HCs |
| Samson et al., 2011 | 21477817 | 21/7 | 12/4 | 41.52 ± 9.76 | 35.83 ± 11.38 | HAMD: 21.29 ± 5.42 | 4.54 ± 5.08 | Unknown | P < 0.05  FEW corrected | No | SPM5 | MNI | Negative valence during passive scene viewing | Negative valence | Potential threat | Sad images vs. netural: MDD > HCs |
| Skokauskas et al., 2015 | 26114449 | 37/25 | 43/23 | 42.27 ± 11.00 | 36.16 ± 12.00 | HAMD: 28.18 ± 6.10 | Unknown | Unknown | P < 0.05  FEW corrected | Unknown | SPM8 | MNI | Positive valence during passive scene viewing | Positive valence | Approach motivation | Judgment of positive emotion vs. judgment of geometrical trial: MDD < HCs; MDD > HCs |
| Studies using experiments mapping the RDoC social processes | | | | | | | | | | | | | | | | |
| Finlayson-Short et al., 2021 | 34215143 | 17/9 | 76/50 | 19.76 ± 2.41 | 21.49 ± 2.20 | MADRS: 25.47 ± 7.20 | Unknown | Unknown | P < 0.05  FDR corrected | No | SPM12 | MNI | Tasks involving processing of self-referential information | Social processes | Perception and understanding of self | Self-referential processing vs. neutral processing: MDD < HCs |
| Suffel et al., 2020 | 32432387 | 33/15 | 43/12 | 37.24 ± 12.33 | 36.47 ± 11.21 | HAMD: 13.17 ± 6.13 | Unknown | Unknown | P < 0.05  corrected | Unknown | SPM12 | MNI | Various facial affect processing tasks | Social process | Social communication | Iconic gesture vs. no gesture: MDD > HCs; MDD < HCs |
| Katayama et al., 2019 | 31170685 | 23/16 | 23/14 | 36.7 ± 9.7 | 40.0 ± 11.3 | HAMD: 21.3 ± 5.2 | Unknown | 1.9 ± 1.0 | P < 0.001  uncorrected | Yes | SPM12 | MNI | Future-thinking-related task | Social processes | Perception and understanding of self | future-thinking-related activation: MDD > HCs; MDD < HCs |
| Komulainen et al., 2018 | 29747140 | 15/9 | 15/- | 23 | Unknown | MADRS: 24.5 ± 4.05; BDI: 31.3 ± 6.73 | Unknown | Unknown | P < 0.05  FEW corrected | No | SPM8 | MNI | Various tasks involving processing of self-referential information | Social processes | Perception and understanding of self | Positive vs. negative self-referential: MDD < HCs |
| Briceno et al., 2013 | 23298715 | 24/24 | 22/22 | 37.8 ± 14.5 | 31.7 ± 14.4 | HDRS: 15.8 ± 7.2 | Unknown | Unknown | P < 0.05  FDR corrected | Yes | SPM2 | Tal | Unspecified valence during facial affect processing | Social process | Social communication | Emotion discrimination vs. animal discrimination: MDD < HCs; MDD > HCs |
| Fournier et al., 2013 | 22571805 | 26/18 | 28/16 | 30.6 ± 7.8 | 32.6 ± 6.4 | HAMD: 21.4 ± 3.9 | 12.2 ± 7.4 | Unknown | P < 0.001  uncorrected | Yes | SPM8 | Tal | Unspecified valence during facial affect processing | Social process | Social communication | All four emotion vs. shapes: MDD > HCs |
| Hao et al., 2015 | 25487911 | 20/6 | 22/9 | 32.3 ± 8.4 | 29.7 ± 5.7 | HAMD: 20.3 ± 7.8 | 2.72/- | Unknown | P < 0.05  AlphaSim corrected | Unknown | SPM8 | MNI | Self-referential processing task | Social Process | Perception and understanding of self (Self) | Non self-serving vs. self-serving: MDD > HCs |
| Regenbogen et al., 2015 | 25573396 | 24/- | 24/- | 36.42 ± 12.01 | 35.25 ± 9.80 | HAMD: 14.45 ± 6.67 | Unknown | Unknown | P < 0.005 corrected | Yes | SPM8 | MNI | Unspecified valence during facial affect processing | Social process | Social communication | Trimodal emotional vs. trimodal neutral: MDD < HCs; MDD > HCs |
| Sarsam et al., 2013 | 24205330 | 13/10 | 14/8 | 32.7 ± 7.6 | 26.4 ± 9.5 | BDI: 29.1 ± 12.9 | Unknown | Unknown | P < 0.005  uncorrected | No | Other | Tal | Self-referential processing task | Social process | Perception and understanding of self (self) | Self vs queen: MDD > HCs |
| Shi et al., 2015 | 26322003 | 29/18 | 33/17 | 20.45 ± 1.80 | 20.75 ± 1.50 | CES-D: 56.10 ± 5.73 | Unknown | Unknown | P < 0.05 FWE  corrected | No | SPM8 | MNI | Unspecified valence during facial affect processing | Social process | Social communication | Emotional faces vs. baseline: MDD > HCs |
| Wagner et al., 2015 | 25872899 | 20/11 | 20/12 | 39.9 ± 13.3 | 34.1 ± 11.3 | HAMD: 23.1 ± 4.6 | Unknown | Unknown | P < 0.05  FWE  corrected | No | SPM8 | Tal | Self-referential processing task | Social process | Perception and understanding of self (self) | Negative self-referential processing vs. baseline: MDD < HCs |
| Whalley et al., 2012 | 22386970 | 15/11 | 15/7 | 33.9 ± 7.7 | 33.8 ± 7.7 | BDI: 29.53 ± 7.87 | Unknown | Unknown | P < 0.05  corrected | Yes | SPM5 | MNI | Self-referential memory paradigm | Social process | Perception and understanding of self (self) | OwnHit vs. elseHit (combination of words and sentences tasks): MDD < HCs |
| Young et al., 2012 | 21798113 | 12/4 | 14/7 | 34 ± 11.0 | 29 ± 9.4 | HAMD: 21 ± 8.30 | Unknown | Unknown | P < 0.001  uncorrected | No | SPM5 | MNI | Self-referential memory paradigm | Social process | Perception and understanding of self (self) | Any memory vs. substraction: MDD < HCs |
| Studies using experiments mapping the RDoC cognitive systems domain | | | | | | | | | | | | | | | | |
| Backes et al., 2014 | 24557502 | 33/20 | 33/20 | 35.79 ± 10.53 | 33.15 ± 12.84 | BDI: 27.27 ± 8.27 | Unknown | Unknown | P < 0.001  corrected | Yes | SPM8 | MNI | Verbal fluency task | Cognitive system | Language | Word generation vs. baseline: MDD > HCs |
| Dietsche et al., 2014 | 24639328 | 23/14 | 23/14 | 36.69 ± 10.35 | 37.13 ± 10.42 | HAMD: 17.75 ± 3.64 | Unknown | Unknown | P < 0.001  corrected | Yes | SPM8 | MNI | Episodic memory encoding/retrieval task | Cognitive system | Declarative memory | Encoding and recognition vs. baseline: MDD < HCs; MDD > HCs |
| Garrett et al., 2011 | 21078708 | 15/10 | 19/6 | 39.81 ± 12.74 | 34.85 ± 12.54 | HAMD: 24.2 ± 3.3 | Unknown | Unknown | P < 0.05  corrected | Yes | SPM5 | Tal | N-back task | Cognitive system | Working memory | 2-back task vs. press for Z: MDD < HCs; MDD > HCs |
| Harvey et al., 2005 | 15955496 | 10/7 | 10/5 | 33.8 ± 8.4 | 29.0 ± 10.1 | MADRS: 26.7 ± 4.6 | 9.7 ± 7.1 | 2.6 ± 1.1 | P < 0.05  corrected | Yes | SPM99 | Tal | N-back task | Cognitive system | Working memory | Nback (1-2-3-back) vs 0-back: MDD > HCs |
| Hugdahl et al., 2007 | - | 9/5 | 12/7 | 36/- | 36/- | MADRS: 26.7 ± 4.6 | Unknown | Unknown | P < 0.001  uncorrected | Yes | SPM99 | MNI | Mental arithmetic task | Cognitive system | Unclassified | Mental arithmetic task vs. vigilance task: MDD < HCs |
| Kassel et al., 2016 | 26831638 | 42/23 | 40/19 | 43.3 ± 16.7 | 37.7 ± 17.3 | HAMD: 15.1 ± 6.4 | Unknown | Unknown | P < 0.05  corrected | Yes | SPM8 | Tal | Semantic list learning task | Cognitive system | Declarative memory | Encoding vs. silent rehearsal and words recalled vs. baseline: MDD < HCs |
| Langenecker et al., 2007 | 17585888 | 22/15 | 22/14 | 41.0 ± 12.2 | 34.2 ± 11.0 | HAMD: 20.4 ± 7.6 | 13.1/- | Unknown | P < 0.0001  uncorrected | No | SPM2 | Unknnown | Go/no-go task | Cognitive system | Cognitive control | Activation for rejections vs. baseline: MDD > HCs |
| Matsuo et al., 2007 | 16983390 | 15/10 | 15/9 | 34.3 ± 11.5 | 37.7 ± 12.1 | HAMD: 20.3 ± 5.3 | 15.3 ± 9.7 | 11.0 ± 9.6 | P < 0.01  corrected | No | FSL | Tal | N-back task | Cognitive system | Working memory | 2-back vs. 1-back: MDD > HCs |
| Matthews et al., 2009 | 19239982 | 15/12 | 16/10 | 24.5/- | 24.3/- | Unknown | Unknown | Unknown | P < 0.05  corrected | No | AFNI | Tal | Go/no-go task | Cognitive system | Cognitive control | Inhibitory processing (hard vs. easy trials): MDD > HCs |
| Naismith et al., 2010 | 20219248 | 19/14 | 20/14 | 56.1 ± 9.8 | 50.6 ± 11.9 | HAMD: 21.6 ± 4.2 | 16.0/- | Unknown | P < 0.005  uncorrected | Yes | FSL | MNI | Implicit learning task | Cognitive system | Unclassified | Implicit learning vs. baseline condition: MDD < HCs; MDD > HCs |
| Remijnse et al., 2013 | 23637737 | 19/7 | 29/20 | 35/- | 33/- | HAMD: 20.1 ± 4.4 | Unknown | Unknown | P < 0.001  uncorrected | No | SPM5 | MNI | Task switching paradigm | Cognitive system | Cognitive control | Switch vs. repeat trials: MDD < HCs |
| Walsh et al., 2007 | 17601497 | 20/14 | 20/13 | 43.7 ± 8.6 | 43.7 ± 8.3 | HAMD: 21.2 ± 2.4 | Unknown | Unknown | P < 0.001  uncorrected | No | Other | Tal | N-back task | Cognitive system | Working memory | 1-2-3-back vs. 0-back: MDD < HCs |
| Walter et al., 2007 | 17197035 | 12/4 | 17/8 | 37.2 ± 9.0 | 30.9 ± 8.8 | HAMD: 18.2 ± 3.7 | 3.1 ± 2.9 | Unknown | P < 0.001  uncorrected | Yes | SPM2 | Tal | Modified sternberg task | Cognitive system | Working memory | All loads vs. control: MDD < HCs; MDD > HCs |
| Werner et al., 2009 | 19346000 | 11/- | 11/- | 37.18 ± 10.35 | 36.27 ± 6.48 | BDI: 20.27 ± 8.74 | Unknown | Unknown | P < 0.05 FDR  corrected | Yes | Other | Tal | Paired associates learning task | Cognitive system | Declarative memory | Encoding and retrieval vs. head templates: |
| Rao et al., 2015 | 26030776 | 16/- | 18/- | 26.4/- | 21.8/- | HAMD: 15/- | 9.7 ± 5.9 | Unknown | P < 0.05  corrected | Yes | SPM8 | MNI | Go/no-go task | Cognitive system | Cognitive control | Correct hits vs. baseline: MDD > HCs |
| Rodriguez-Can et al., 2014 | 25066663 | 26/16 | 52/32 | 46.50 ± 13.29 | 46.25 ± 10.21 | HAMD: 26.84 ± 4.48 | 10.96 ± 10.54 | Unknown | P < 0.05 FWE  corrected | Yes | FSL | MNI | N-back task | Cognitive system | Working memory | 2-back vs. baseline: MDD < HCs |
| Rose et al., 2006 | 16157491 | 9/- | 9/- | 32.00 ± 7.86 | 31.33 ± 8.32 | HAMD: 20.56 ± 5.59 | 4.35 ± 3.79 | 1.9 ± 1.4 | P < 0.05  corrected | Yes | SPM99 | MNI | N-back task | Cognitive system | Working memory | Linear increase in the level of difficulty (3-2-1-back vs. 0-back): MDD > HCs |
| Takamura et al., 2016 | 28052303 | 16/6 | 16/6 | 39.4 ± 9.1 | 37.3 ± 11.9 | HAMD: 20.7 ± 5.5 | Unknown | Unknown | P < 0.05 FWE  corrected | Yes | SPM8 | MNI | Verbal fluency task | Cognitive system | Language | Semantic verbal fluency task vs. baseline: MDD < HCs; MDD > HCs |
| Young et al., 2014 | 25065602 | 16/10 | 16/10 | 34.2 ± 9.06 | 27.3 ± 8.02 | HAMD: 20.0 ± 6.82 | Unknown | Unknown | P < 0.05  corrected | No | AFNI | Tal | Episodic memory retrieval task | Cognitive Systems | Memory | Positive or negative Specific memory vs. Positive or negative Example Generation: MDD < HCs; MDD > HCs |
| Barch et al., 2003 | 12614990 | 14/7 | 49/27 | 37.6 ± 12.0 | 36.5 ± 11.2 | HAMD: 21.7 ± 6.1 | Unknown | Unknown | P < 0.05  corrected | Yes | Other | Tal | N-back task | Cognitive system | Working memory | 2-back vs. baseline: MDD < HCs |
| Dai et al., 2018 | 29575563 | 30/23 | 33/21 | 48.77 ± 11.88 | 48.85 ± 12.86 | HDRS: 23.43 ± 2.94 | 8.75 ± 6.94 | 3.1 ± 1.37 | P < 0.05  FEW corrected | No | SPM8 | MNI | Inhibition of return task | Cognitive systems | Cognitive Control | Invalid cue vs. valid cue:  MDD > HCs |
| Yüksel et al., 2018 | 29421266 | 74/41 | 74/41 | 36.8 ± 10.7 | 35.8 ± 10.8 | BDI: 22.9 ± 10.9 | Unknown | Unknown | P < 0.01  corrected | Yes | SPM8 | MNI | N-back tasks | Cognitive systems | Working memory | 2-back vs. 0-back: MDD < HCs;  3-back vs. 0-back: MDD < HCs;  2-back vs. 3-back: MDD < HCs |
| Ai et al., 2018 | 30212727 | 103/64 | 26/13 | 37.20/- | 38.96 ± 7.94 | IDS: 25.58/- | Unknown | Unknown | P < 0.05 FWE  corrected | Yes | SPM8 | MNI | Tower of London | Cognitive systems | Cognitive control | Planning vs. baseline: MDD < HCs |
| Rodríguez-Cano et al., 2017 | 28714580 | 26/- | 26/- | 46.50 ± 13.30 | 46.77 ± 11.18 | HDRS: 27.00 ± 4.51 | 10.50 ± 9.78 | 3.69 ± 3.77 | P < 0.05  FEW corrected | Yes | FSL | MNI | N-back tasks | Cognitive systems | Working memory | N-back (1-2-back) vs. baseline: MDD < HCs; MDD > HCs |
| Sacchet et al., 2017 | 27649971 | 16/9 | 16/8 | 31.5 ± 8.9 | 31.7 ± 10.0 | BDI: 31.1 ± 9.5 | Unknown | Unknown | P < 0.005  uncorrected | Yes | AFNI | Tal | Implicit learning tasks | Cognitive systems | Cognitive unclassified tasks | No think vs. think: MDD > HCs |
| Le et al., 2017 | 28138426 | 18/12 | 21/12 | 22.0 ± 3.09 | 22.19 ± 3.38 | IDS: 40.79 ± 8.70 | 3.87 ± 2.61 | Unknown | P < 0.05  FDR corrected | No | SPM8 | MNI | Postcue delay of the working memory task | Cognitive systems | Working memory | Remember scene vs. remember face: MDD < HCs |
| Tavares et al., 2008 | 18586109 | 13/10 | 15/11 | 38.3 ± 2.3 | 33.9 ± 2.33 | MADRS: 23 ± 1.7 | Unknown | Unknown | P < 0.05  corrected | No | SPM2 | MNI | Reversal learning task | Cognitive system | Cognitive control | Perform tasks vs. baseline: MDD < HCs |
| Crane et al., 2016 | 27454009 | 18/11 | 54/38 | 34.28 ± 11.69 | 33.80 ± 11.56 | HDRS:20.76 ± 7.25 | Unknown | Unknown | P < 0.005  corrected | No | SPM8 | MNI | Go/No-Go task | Cognitive system | Cognitive control | Activation for targets vs. baseline: MDD < HCs |
| Study including cross-domain experiments | | | | | | | | | | | | | | | | |
| Cerullo et al., 2014  n/p | 24990479 | 25/17 | 25/17 | 27 ± 7 | 26 ± 7 | HAMD: 38 ± 8 | Unknown | Unknown | P < 0.05  corrected | No | AFNI | Tal | Emotional visual oddball task | Cognitive affective interaction | Cognitive affective interaction | Emotional images, circles vs. square trials: MDD < HCs; MDD > HCs |
| Chechko et al., 2013 | 23394712 | 18/13 | 18/13 | 36.5 ± 10.8 | 36.0 ± 10.3 | HAMD: 22.7 ± 5 | 6.2/- | Unknown | P < 0.05 FWE  corrected | Yes | SPM5 | MNI | Emotional stroop task | Cognitive affective interaction | Cognitive affective interaction | Incongruent vs. congruent in emotional interference: MDD < HCs |
| Dichter et al., 2009 | 18706701 | 14/7 | 15/9 | 34.8 ± 14.3 | 30.8 ± 9.6 | BDI: 26.9 ± 4.9 | Unknown | Unknown | P < 0.05  corrected | No | FSL | MNI | Emotional visual oddball task | Cognitive affective interaction | Cognitive affective interaction | Targets within sad blocks vs. baseline: MDD < HCs |
| Greening et al., 2009 | 23482626 | 19/13 | 19/13 | 26.79 ± 11.4 | 27.63 ± 11.0 | BDI: 25.53 ± 10.4 | Unknown | Unknown | P < 0.05 FWE  corrected | No | Other | MNI | Emotion regulation task | Cognitive affective interaction | Cognitive affective interaction | Reduce vs. attend: MDD > HCs |
| Heller et al., 2009 | 20080793 | 27/15 | 19/10 | 31.48 ± 11.58 | 31.84 ± 14.65 | HAMD: 20.6 ± 2.39 | Unknown | Unknown | P < 0.05  corrected | No | AFNI | Tal | Emotion regulation task | Cognitive affective interaction | Cognitive affective interaction | Enhance vs. suppress condition: MDD < HCs |
| Johnstone et al., 2007 | 17699669 | 21/13 | 18/11 | 33 ± 12 | 28 ± 12 | HAMD: 21 ± 2.5 | Unknown | Unknown | P < 0.01  corrected | No | AFNI | Tal | Emotion regulation task | Cognitive affective interaction | Cognitive affective interaction | Decrease condition vs. attend condition: MDD > HCs |
| Lisiecka et al., 2013 | 23010257 | 50/33 | 46/25 | 42.7/- | 37.35/- | HAMD: 28.5 | 9.6 ± 13.4 | Unknown | P < 0.05 FWE  corrected | Yes | SPM8 | MNI | Emotional attention shifiting task | Cognitive affective interaction | Cognitive affective interaction | Emotion processing, attention shifting vs. baseline: MDD < HCs |
| Mitterschiffthaler et al., 2008 | 17825123 | 17/14 | 17/14 | 39.3 ± 9.4 | 39.4 ± 9.2 | HAMD: 20.88 ± 1.83 | Unknown | Unknown | P < 0.05 FWE  corrected | No | SPM2 | Tal | Negative words vs. neutral words: | Cognitive affective interaction | Cognitive affective interaction | Negative words vs. neutral words: MDD > HCs |
| Wang et al., 2008 | 18455373 | 19/12 | 20/13 | 39.3 ± 9.0 | 36.5 ± 10.5 | HAMD: 19.9 ± 5.3 | 21.6/- | Unknown | P < 0.05 FDR  corrected | Yes | SPM99 | MNI | Emotional visual oddball task | Cognitive affective interaction | Cognitive affective interaction | Sad vs neutral distractors, targets vs. scrambled pictures: MDD > HCs |
| Lemke et al., 2022 | 34102346 | 333/195 | 333/200 | 36.80 ± 10.57 | 36.64 ± 12.49 | HDRS: 19.61 ± 5.88 | 3.24 ± 4.55 | 3.71 ± 4.2 | P < 0.001  uncorrected | Yes | SPM12 | MNI | Emotional identity task | Cross Domain tasks | Cross-domain affective | Negative emotional identity vs. neutral shape identity:  MDD > HCs |
| Alders et al., 2020 | 31400735 | 48/33 | 30/22 | 34.7 ± 12.2 | 33.2 ± 9.8 | MADRS: 30 ± 6 | 29 ± 33 | Unknown | P < 0.05  corrected | Yes | FSL | MNI | Emotional conflict task | Cross domain task | Cross domain | Incongruent vs. congruent: MDD < HCs |
| Amico et al., 2012 | 22738278 | 14/9 | 14/4 | 41.2 ± 10.3 | 35.0 ± 9.4 | HDRS: 24.8 ± 5.0 | 16 ± 10 | Unknown | P < 0.05  FEW corrected | Yes | SPM8 | MNI | Emotional attention shifting task | Cross Domain tasks | Cross Domain | Dot-probe task using fearful stimuli vs. neutral stimuli: MDD < HCs |
| Avery et al., 2014 | 24387823 | 20/13 | 20/12 | 36 ± 9 | 33 ± 7 | HDRS: 23.1 ± 7.5 | 5.08 ± 6.25 | Unknown | P < 0.05  corrected | No | AFNI | Tal | Sensation odd task | Cognitive systems | Attention | Interoception vs. exteroceptive attention: MDD < HCs |
| Canli et al., 2004 | 15570157 | 15/12 | 15/12 | 35.1/- | 30.7/- | BDI: 23.9 ± 7.53 | Unknown | Unknown | P < 0.001  uncorrected | Yes | SPM99 | Tal | Positive valence during passive word | Positive valence | Approach motivation | Happy vs. neutral: MDD < HCs |
|  |  |  |  |  |  |  |  |  |  |  |  |  | Negative valence during passive word | Negative valence | Potential threat | Sad, social threat and physical threat vs. neutral: MDD < HCs; MDD > HCs |
| Chechko et al., 2016 | 27672555 | 21/16 | 24/14 | 36.5 ± 10.8 | 26.6 ± 3.2 | HAMD: 22.9 ± 4.8 | Unknown | Unknown | P < 0.05  FEW corrected | Yes | SPM5 | MNI | Emotional interference task | Cross Domain tasks | Cross Domain | Incongruent vs. congruent: MDD < HCs |
| Gaebler et al., 2020  n/p | 31220318 | 27/8 | 31/14 | 37.4 ± 10.4 | 36.4 ± 10.0 | BDI: 23.9 ± 9.8 | Unknown | Unknown | P < 0.001  corrected | Yes | Other | Tal | Auditory mismatch processing task | Cross Domain tasks | Cross Domain | Auditory mismatch blocks vs. baseline: MDD < HCs |
| Ma et al., 2021 | 34305677 | 37/23 | 54/29 | 25.89 ± 4.75 | 23.94 ± 3.05 | HAMD: 23.35 ± 5.60 | 1.52 ± 2.49 | Unknown | P < 0.05  FEW corrected | Yes | SPM12 | MNI | Working memory task under stress | Cross Domain tasks | Cross Domain | Working memory under stress vs. baseline: MDD < HCs |
|  |  |  |  |  |  |  |  |  |  |  |  |  | Working memory task | Cognitive systems | Working memory | Working memory task vs. baseline: MDD < HCs |
| Hall et al., 2014 | 23990079 | 29/16 | 29/16 | 37.38/- | 37.69/- | HAMD: 13.52/- | 16.09 ± 8.80? | 5.47 ± 5.95 | P < 0.005  corrected | Yes | Other | Tal | Contingency reversal reward task | Cross Domain tasks | Cross Domain | Reward acquisition vs. punishment reversal: MDD < HCs; MDD > HCs |
|  |  |  |  |  |  |  |  |  |  |  |  |  | Reward task | Positive valence | Reward attainment | Magnitude of reward: large gain vs. small gain: MDD < HCs |
|  |  |  |  |  |  |  |  |  |  |  |  |  | Punishment task | Negative valence | Frustrative non-reward | Magnitude of punishment: large loss vs. small loss: MDD < HCs |
| Koch et al., 2018 | 29682814 | 16/16 | 14/14 | 29.9 ± 8.9 | 26.8 ± 4.3 | HDRS: 20.9 ± 5.1 | Unknown | Unknown | P < 0.05  FEW corrected | Yes | SPM12 | MNI | Emotional identity task | Cross domain tasks | Cross domain | Emotional vs. neutral prosody: MDD > HCs |
| Scott et al., 2015 | 25335914 | 16/13 | 16/13 | 39.94 ± 9.48 | 40.00 ± 9.27 | HAMD: 20.88 ± 1.89 | 1.64/- | 0.63/- | P < 0.05 corrected | No | Other | Tal | Statements from a modified dysfunctional attitudes task | Cross Domain tasks | Cross Domain | Extreme attributions in dysfunctional attitudes scale vs. baseline: MDD < HCs; MDD > HCs |
| DeVille et al., 2019 | 29724684 | 24/15 | 21/12 | 29.3 ± 8.0 | 30．8 ± 10.0 | HAMD: 22.5 ± 6.59 | Unknown | Unknown | P < 0.0005  uncorrected | No | AFNI | Tal | Interoceptive encoding and recall task | Cross domain tasks | Cross domain | Interoceptive recall vs. exteroceptive recall: MDD < HCs |
| Wang et al., 2018 | 29163097 | 12/7 | 15/8 | 29.50 ± 8.46 | 25.80 ± 5.89 | HAMD: 25.17 ± 5.18 | Unknown | Unknown | P < 0.05 AlphaSim-corrected | No | SPM8 | MNI | Emotion regulation task | Cross domain tasks | Cross domain | Emotional reappraisal vs. emotional attend: MDD > HCs; MDD < HCs |
| Bermpohl et al., 2009 | 19632301 | 15/12 | 21/10 | 43.4 ± 10.2 | 24.1 ± 2.5 | HDRS: 24.7 ± 3.6 | Unknown | Unknown | P < 0.001  uncorrected | Yes | SPM2 | MNI | Unspecified valence during passive scene viewing | Cross-domain affective | Cross-domain affective | Emotional vs. neutral expectancy: MDD < HCs; MDD > HCs |
| Grimm et al., 2008 | 17888408 | 29/21 | 19/11 | 35.32 ± 7.26 | 40.00 ± 9.89 | HAMD: 33.12 ± 7.13 | 6.6 ± 8.1 | 1.8 ± 2.2 | P < 0.001  uncorrected | No | SPM2 | MNI | Unspecified valence during passive scene viewing | Cross-domain | Cross-domain | Emotional judgment vs. passive viewing: MDD < HCs; MDD > HCs |
| Ritchey et al., 2011 | 20934190 | 22/13 | 14/9 | 36.1 ± 10.1 | 34.6 ± 6.9 | HAMD: 26.7 ± 6.7 | Unknown | Unknown | P < 0.005  uncorrected | No | SPM2 | Tal | Unspecified valence during passive scene viewing | Cross-domain | Cross-domain | Negative & positive vs. neutral: MDD < HCs |
| Tozzi et al., 2016 | 26076833 | 40/27 | 43/27 | 36.15/- | 41.58/- | HAMD: 28.4/- | 14.69 | 27.24 | P < 0.05  FWE  corrected | Yes | SPM12 | MNI | Unspecified valence during passive scene viewing | Cross-domain | Cross-domain | Emotional vs geometrical: MDD < HCs |
| Tremblay et al., 2005 | 16275810 | 12/6 | 12/7 | 34.83 ± 13.96 | 29.33 ± 9.31 | HAMD: 27.75 ± 3.05 | Unknown | Unknown | P < 0.001  uncorrected | No | Other | Tal | Unspecified valence during passive scene viewing | Cross-domain | Cross-domain | All pictures vs. fixation crosses: MDD > HCs |
| Victor et al., 2010 | 21041614 | 22/12 | 25/15 | 31 ± 7.8 | 28.8 ± 6.7 | HAMD: 24 ± 6.3 | Unknown | Unknown | P < 0.001  uncorrected | Yes | SPM5 | Tal | Unspecified valence during passive scene viewing | Cross-domain | Cross-domain | Unmasked-sad vs. unmasked-happy faces: MDD < HCs; MDD > HCs |

MADRS, Montgomery-Åsberg Depression Rating Scale; IDS, Inventory of depressive symptomatology; BDI, Beck Depression Inventory; HAMD: Hamilton Depression Scale; HDRS, Hamilton Depression Rating Scale; HCs, healthy controls; MDD, major depressive disorder; MNI, Montreal Neurological Institute; Tal, Talairach; CES-D: Center for Epidemiologic Studies Depression Scale
